# OCT-based Visual Field Estimation via Segmentation-free 3D CNNs Shows Lower Longitudinal Variability than Standard Automated Perimetry

**DOI:** 10.1101/2024.08.17.24312150

**Authors:** Makoto Koyama, Hidenori Takahashi, Satoru Inoda, Chihiro Mayama, Yuta Ueno, Yoshikazu Ito, Tetsuro Oshika, Masaki Tanito

## Abstract

**Purpose:** To train and evaluate segmentation-free 3D convolutional neural network (3DCNN) models for estimating visual field (VF) from optical coherence tomography (OCT) images and to independently assess the longitudinal variability and progression detection capabilities of Humphrey Field Analyzer (HFA) measurements and OCT-based estimated VF (OCT-VF) in a diverse clinical population.

**Design:** Retrospective multicenter study.

**Participants:** 13,366 patients (24,313 eyes) underwent HFA tests (24-2, trimmed 30-2, or 10-2 test patterns) and macular OCT imaging at five ophthalmic institutions. The dataset included 129,007 paired OCT-VF data points representing various ocular conditions.

**Methods:** We trained segmentation-free 3DCNN models using comprehensive OCT datasets without disease-specific exclusions, employing 10-fold cross-validation to estimate VF thresholds and mean deviation (MD). Unlike previous studies, we independently assessed both OCT-VF and HFA measurements by creating separate longitudinal datasets with standardized measurement counts and observation periods for comparative analysis, enabling direct evaluation of clinical reliability. We analyzed absolute residual variability from regression lines using jackknife resampling, applied Bonferroni correction for multiple comparisons, and used Spearman’s correlation for progression analysis.

**Main Outcome Measures:** OCT-VF and HFA VF agreement, residual variability, progression detection rates, and progression rate correlations.

**Results:** OCT-VF and HFA VF showed correlations (Pearson’s r: 24-2 thresholds 0.863, MD 0.924; 10-2 thresholds 0.881, MD 0.939; all p < 0.001). OCT-VF demonstrated significantly lower residual variability than HFA for all parameters (OCT-VF vs. HFA: 0.58 vs. 1.12 dB for 24-2 MD; 0.70 vs. 1.12 dB for 10-2 MD; all p < 0.001). This advantage persisted across all test points (mean variability reduction: 60.4% for 24-2; 55.1% for 10-2), age groups, and most severity levels. OCT-VF identified more progression events (24-2 MD: 113% more, 10-2 MD: 48.6% more). MD slopes showed correlations between OCT-VF and HFA (Pearson’s r: 24-2 MD 0.831, 10-2 MD 0.863; all p < 0.001).

**Conclusions:** The segmentation-free 3DCNN models objectively estimated VF from OCT images with significantly lower longitudinal variability than performance-dependent HFA measurements across diverse ocular conditions. The lower variability of OCT-VF enhances statistical power for progression detection, suggesting its clinical potential as a complementary tool derived from routine OCT data, decreasing measurement noise, and enabling more timely therapeutic interventions.

## Introduction

Visual field (VF) testing is crucial for diagnosing and monitoring various ocular conditions, particularly glaucoma, a leading cause of irreversible blindness worldwide.^1–3^ Although the Humphrey Field Analyzer (HFA; Carl Zeiss Meditec, Jena, Germany) remains the gold standard for VF assessment, HFA testing has recognized limitations, including its performance-dependent nature, high test–retest variability, and time-consuming process.^2,4,5^

Optical coherence tomography (OCT) has revolutionized ophthalmic imaging, providing high-resolution, objective, and reproducible assessments of ocular structures.^6^ The established structure-function relationship between OCT-measured retinal parameters and functional deficits observed in VF testing has created opportunities for more objective VF assessment. Against this backdrop, recent advances in artificial intelligence (AI) have enabled the development of approaches that estimate VF directly from OCT images, potentially addressing many limitations of conventional perimetry.^7–15^

Various OCT-based VF estimation models exist, including 2D and 3D approaches. While 2D methods offer computational efficiency, they have limitations: segmentation-based methods reduce generalizability,^7,8^ and B-scan-based methods have partial imaging coverage.^9–11^ We adopted a 3DCNN model because 3D approaches leverage complete volumetric data to capture the full depth and spatial complexity of the retinal architecture, allowing for potentially more accurate estimations, as suggested by previous studies.^12–14^ This volumetric approach preserves the intricate 3D relationships between retinal layers critical for understanding structure-function relationships, which may substantially reduce variability. Our previous research has demonstrated that training on comprehensive datasets, including various ocular conditions, significantly outperforms glaucoma-specific training in estimating VF from 3D OCT images.^15^ Building on these findings, the current study employed comprehensive training on diverse ocular conditions to enhance generalizability and avoid selection bias.

Prior research has typically approached VF estimation by treating HFA as a reference, aiming to predict HFA measurements from OCT data.^7–15^ However, this approach has limitations due to HFA’s inherent response biases and increased test–retest variability with disease severity.^2,4^ Unlike previous studies, we evaluated our model by independently analyzing both methods’ internal consistency and variability rather than merely assessing concordance with HFA measurements. This balanced approach acknowledged the variability inherent in both assessment methods and compared their residual variability over time to determine which provided more consistent and clinically useful information for disease monitoring.

The primary aims of this study were to (1) train and evaluate segmentation-free 3DCNN models for objectively estimating VF from OCT images in a diverse clinical population, (2) compare the residual variability of OCT-based estimated VF (OCT-VF) with that of HFA measurements, and (3) assess the comparative ability of both methods to detect VF progression, potentially offering a more reliable approach for VF assessment and progression monitoring in clinical practice. This research may provide insights into the potential of objective OCT-based VF estimation to complement or enhance conventional performance-dependent perimetry, ultimately improving disease monitoring and patient outcomes in clinical settings. The reduced measurement variability could enable earlier detection of true progression, potentially allowing for more timely therapeutic interventions that preserve visual function.

## Methods

### Study Design and Participants

The Institutional Review Boards of Shimane University Hospital (IRB No. KS20230719-3, approved on Aug 10, 2023) and Jichi Medical University (IRB No. Rindai24-130, approved on Feb 4, 2025) approved this retrospective multicenter study. Participating institutions included Shimane University Hospital, Shimane, Japan; Jichi Medical University, Tochigi, Japan; JCHO Tokyo Shinjuku Medical Center, Tokyo, Japan; Omiyahamada Eye Clinic, Saitama, Japan; and Minamikoyasu Eye Clinic, Chiba, Japan. This study adhered to the tenets of the Declaration of Helsinki. Due to the retrospective nature of the study, the requirement for informed consent was waived by the respective IRBs. An opt-out approach was employed, and study information was publicly available on each institution’s website and premises, allowing patients to decline participation. We retrospectively collected data from patients who underwent macular OCT imaging and HFA testing between Aug 5, 2004, and Feb 12, 2025.

### Inclusion Criteria

We retrospectively included all consecutive eyes that met the following criteria: (1) at least one macular OCT scan with a signal strength index (SSI) ≥7 and (2) at least one VF test using the HFA with a 30-2, 24-2, or 10-2 test pattern using Swedish Interactive Threshold Algorithm (SITA) strategy. To reflect real-world clinical practice, no exclusions were made based on ocular diagnosis, and both SITA-Standard and SITA-Fast strategies were used for model training and evaluation. Peripheral test points from 30-2 VF tests were trimmed to match the 24-2 pattern. Reliability thresholds were set at <33% for all three indices: false-positive rate, false-negative rate, and fixation loss rate. This threshold was based on prior experimentation and is consistent with previous studies.^7,9,15^ OCT images were acquired using RS-3000, RS-3000 Advance, RS-3000 Advance2, or Mirante devices (Nidek, Gamagori, Japan) with a 9 mm × 9 mm macular scan protocol, performed under both dilated and undilated conditions. Since SITA-Standard is generally considered to have lower test–retest variability,^5^ we also conducted SITA-Standard–only analyses for key outcomes, as presented in the Supplementary Materials.

### Data Acquisition and Preprocessing

We paired OCT scans with HFA tests performed within 90 days of the OCT acquisition date, selecting the HFA test with the smallest absolute time difference when multiple tests were available. For eyes with both 24-2 and 10-2 test patterns, we included all available data. For eyes with only one test pattern (e.g., only 24-2 but not 10-2) within the 90-day window, we used mask values to indicate missing data points. We included all eligible paired data from eyes with multiple OCT scans and VF tests in the model development dataset, using the raw thresholds and mean deviation (MD) values without any clipping.

### Model Architecture and Configuration

We adopted a segmentation-free 3DCNN model based on the EfficientNet3D-b0 architecture^16^ with a 30% dropout layer to reduce overfitting (Fig. 1). This architecture was selected for its optimal balance of model complexity and computational efficiency for 3D volumetric data. The model was trained from scratch using a comprehensive OCT dataset. Input OCT images were standardized to a resolution of 224 × 224 × 128, and z-score normalization was applied to both images and VF data. The model outputs estimated VF thresholds (52 points for 24-2 and 68 points for 10-2 patterns) and their respective MDs. Following previous reports,^8,15^ we employed horizontal flipping for left eye data and vertical flipping as data augmentation techniques. Model training utilized the Adam optimizer with a mini-batch size of 4 and a variable learning rate schedule, minimizing mean squared error while accounting for missing data points through masked backpropagation.

**Figure 1.**
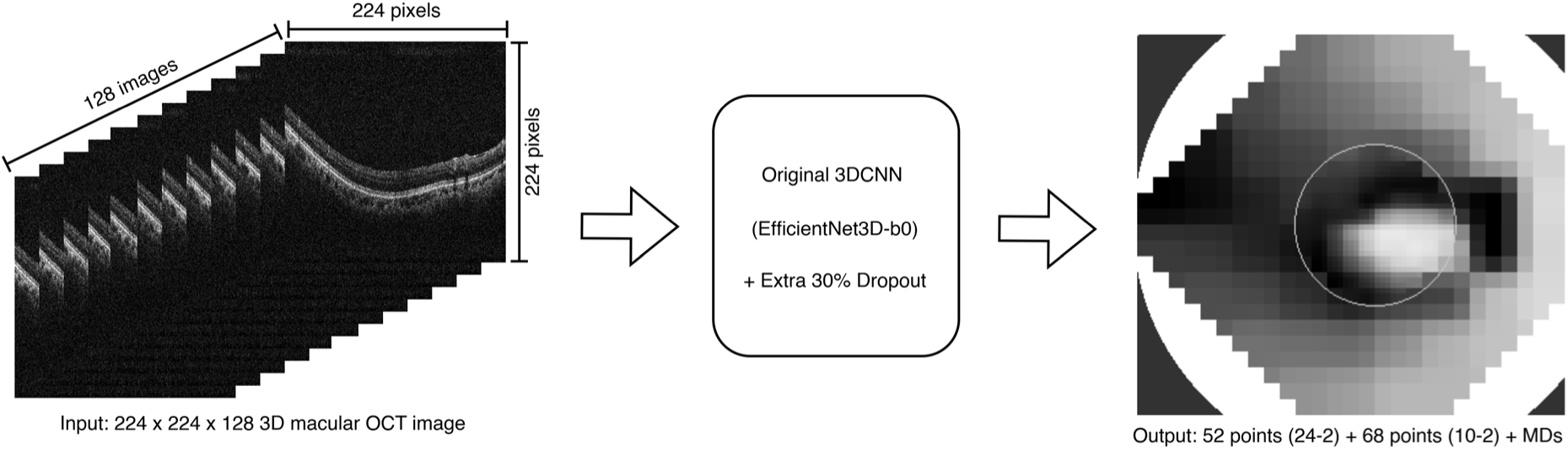
Schematic representation of the segmentation-free 3DCNN model. We based the model on the EfficientNet3D-b0 architecture with an additional 30% dropout layer, which directly connects to the output layer and 122 outputs: VF thresholds (52 points for 24-2 and 68 points for 10-2) and their respective MD values. 3DCNN = 3D convolutional neural network; OCT = optical coherence tomography; VF = visual field; MD = mean deviation.

### Model Training and Evaluation

We employed a 10-fold cross-validation approach using stratified sampling to ensure balanced distribution across folds. Patients were stratified by institution and further divided into 16 subgroups based on test pattern (24-2 or 10-2), MD value (above or below the mean), and number of measurements per eye (above or below the mean). This stratified, patient-wise splitting ensured that data from the same patient did not appear in more than one set, thereby preventing data leakage while maintaining representative distributions of disease severity and measurement frequency across the training, validation, and test sets (8:1:1 ratio). For evaluation, we selected the epoch showing the best performance on the validation set and used the corresponding model weights to assess performance on the test set. During post-processing, any OCT-VF threshold values below 0 dB were set to 0 dB.

We calculated the mean absolute error (MAE), Pearson’s and Spearman’s correlation coefficients, and the intraclass correlation coefficient (ICC(2,1)) to assess the relationships between OCT-VF and HFA parameters. We used Bland-Altman plots to evaluate the agreement between OCT-VF and HFA MDs. Additionally, we analyzed the relationships between the MAE and VF severity, refractive errors, and individual test points to assess the model’s performance across different clinical scenarios and VF regions.

### Independent Longitudinal Analysis for Comparative Reliability Assessment

Following the model performance evaluation, we conducted a separate analysis to compare the longitudinal reliability of OCT-VF and HFA measurements by creating independent datasets for both methods, without disease-based exclusions. For the OCT-VF dataset, we generated estimates from all available macular OCT images (SSI ≥7) using our trained 3DCNN models. This dataset was distinct from the original paired OCT-HFA dataset used for model training and initial evaluation. We assigned each patient to the appropriate model based on their test fold allocation in the cross-validation process, with patients not included in any fold assigned to a randomly selected model. For the HFA dataset, we included all HFA tests, regardless of their false-positive, false-negative, or fixation loss rates, to reflect real-world clinical conditions. We used the thresholds and MD values without any clipping for both groups, except OCT-VF threshold values below 0 dB were set to 0 dB.

To minimize within-subject variability, we averaged multiple VF measurements taken on the same day at each test point in each group separately. Then, all thresholds in both groups were rounded to integers to maintain consistency with the original HFA measurement scale. To align both groups temporally, we defined a valid period common to both groups from the earliest to the latest measurement date, discarding measurements outside this period. When measurements were not taken on the same day in both groups, we extended the valid period to include the nearest available value. We included only eyes with at least six measurements in both groups, excluding any eyes that did not meet this threshold. Furthermore, for each eye, we identified the group with fewer measurements and trimmed the excess measurements from the other group to achieve measurement uniformity by dividing the observation period into equal intervals and selecting the measurements closest to each interval boundary.

### Statistical Analysis

We assessed residual variability using jackknife resampling by iteratively excluding data points, fitting regression lines, and calculating absolute residuals—a technique selected for its robustness to outliers and suitability for dependent longitudinal measurements. We compared OCT-VF and HFA datasets using generalized estimating equations (GEE), adjusting for follow-up duration, age, clustering by eye and patient, and measurement point location for threshold analyses only. We independently evaluated OCT-VF and HFA datasets for longitudinal analysis to avoid biases from performance-dependent testing.

For progression analysis, we divided VF into anatomically relevant clusters (Fig. 2): 10 clusters for the 24-2 pattern based on the Glaucoma Hemifield Test,^17^ and 8 clusters for the 10-2 pattern based on Suzumura et al.’s classification.^18^ We analyzed longitudinal correlations using Spearman’s rank correlation coefficient and defined significant progression as cases with a statistically significant p-value derived from Spearman’s rank correlation test with a negative correlation coefficient. To address multiple comparisons, we applied Bonferroni correction (p=0.0125) for the four primary outcomes and Holm’s method for cluster-specific analyses. We performed all analyses using Python 3.12.9 with scikit-learn (1.6.1), statsmodels (0.14.4), and PyTorch (2.6.0).

**Figure 2.**
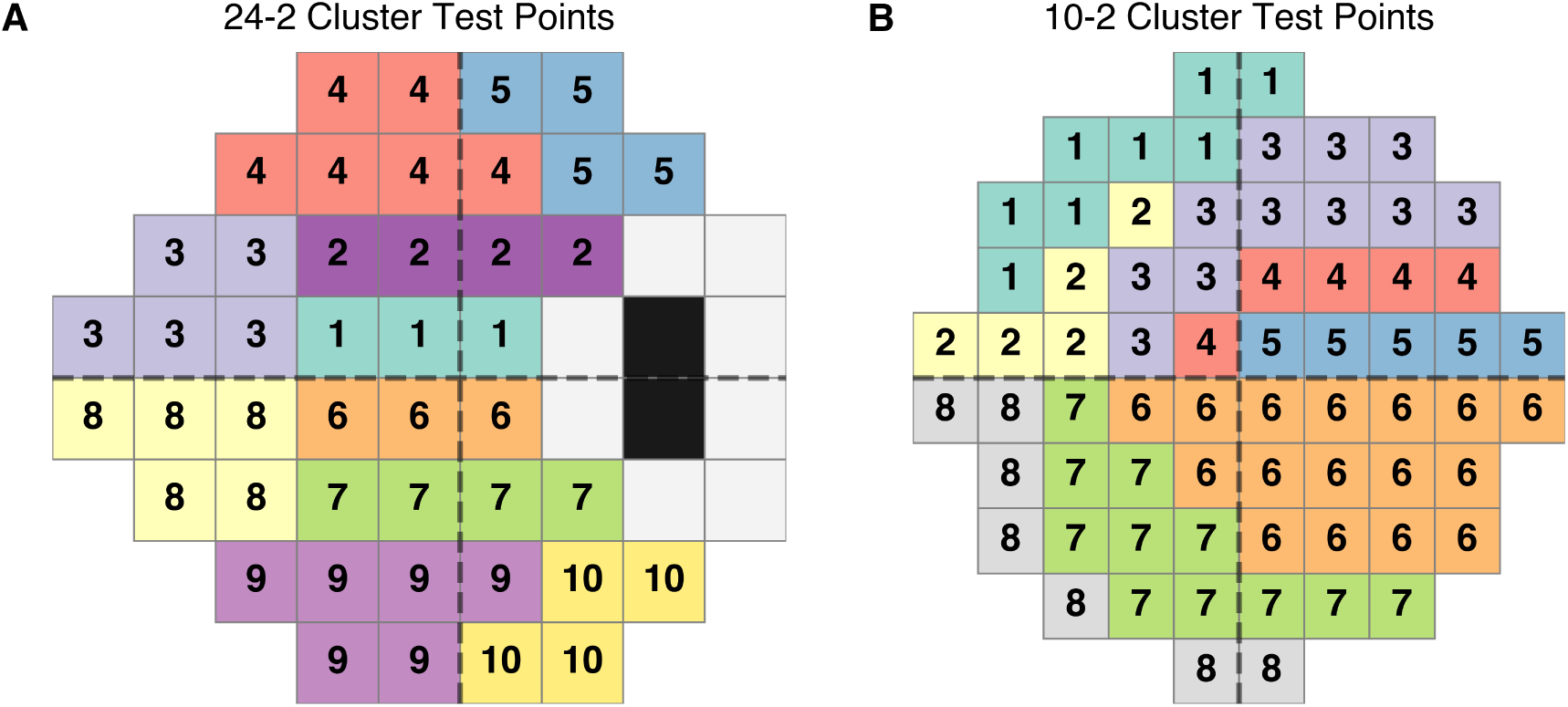
Visual field test point clusters used for progression analysis. (A) 24-2 test pattern with 10 clusters based on the Glaucoma Hemifield Test regions. (B) 10-2 test pattern with 8 clusters based on Suzumura et al.’s classification. Different colors represent different cluster regions used for statistical analysis of progression.

## Results

### Model Performance in Estimating Visual Field

Table 1 provides an overview of the population characteristics for the dataset used to train the 3DCNN models, including the number of patients, eyes, and paired OCT-VF data points used during model development. Pearson’s correlation coefficients (r) for VF thresholds and MDs were 24-2 thresholds r=0.863, MD r=0.924; 10-2 thresholds r=0.881, MD r=0.939 (all p < 0.001, Table 2 and Fig. 3). The Bland-Altman analysis showed mean differences between OCT-VF and HFA values less than -0.12 dB for all analyses, with correlation coefficients ranging from -0.28 to -0.13 (Fig. 4).

**Figure 3.**
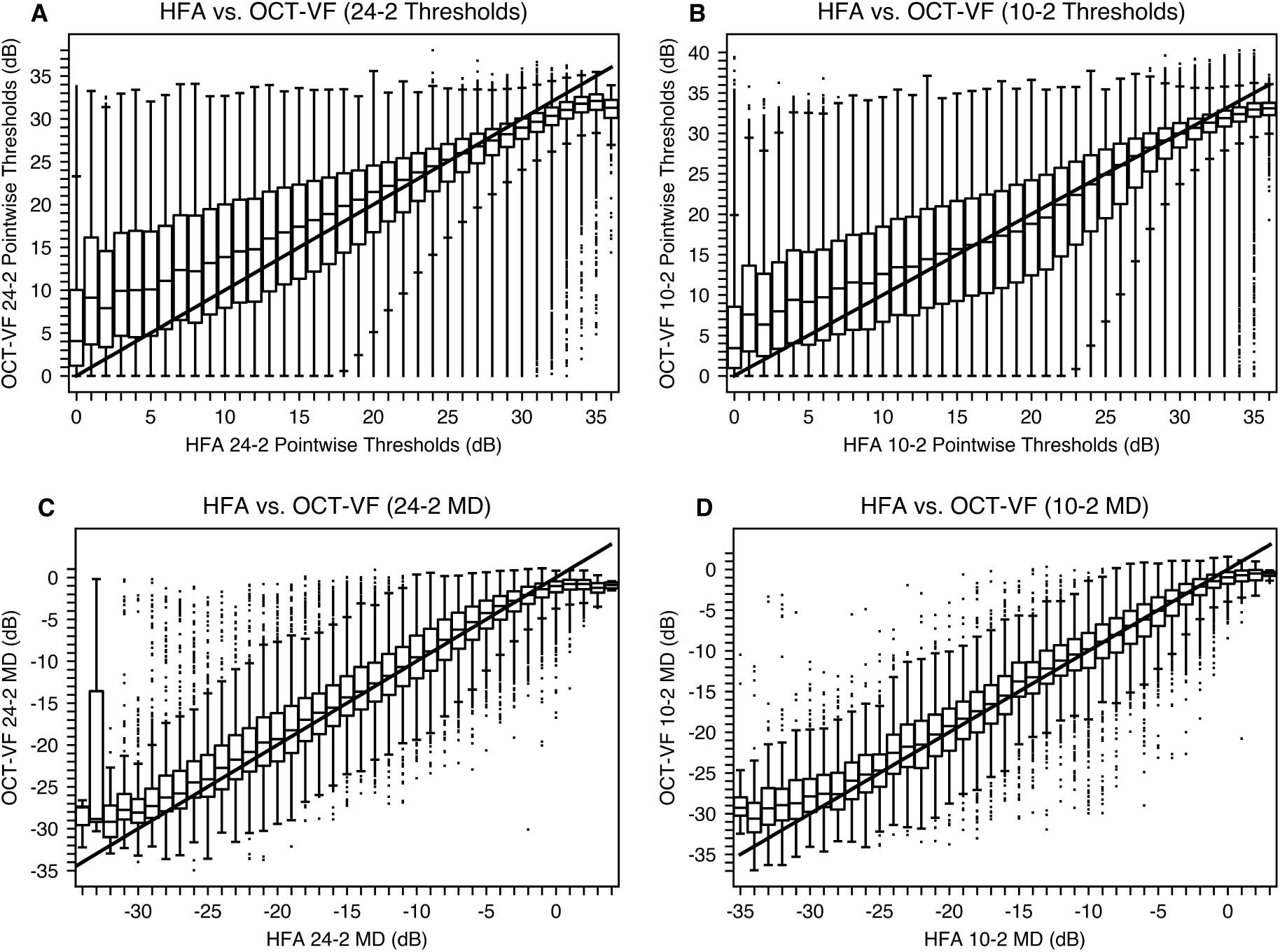
Correlations between HFA and OCT-VF parameters. (A) 24-2 VF threshold (pointwise values, Pearson’s r = 0.863), (B) 10-2 VF threshold (pointwise values, Pearson’s r = 0.881), (C) 24-2 MD (Pearson’s r = 0.924), and (D) 10-2 MD (Pearson’s r = 0.939). All correlations are significant at p < 0.001. Each panel displays boxplots of estimated values for each measured value and an identity line (x=y). Note the increased error in panel (C) at the MD value of -33 dB. Preliminary investigations from a single-center subset suggested that these outliers were primarily associated with non-organic VF loss, testing errors, or intracranial pathologies such as bitemporal hemianopia. OCT = optical coherence tomography; VF = visual field; OCT-VF = OCT-based estimated visual field; MD = mean deviation.

**Figure 4.**
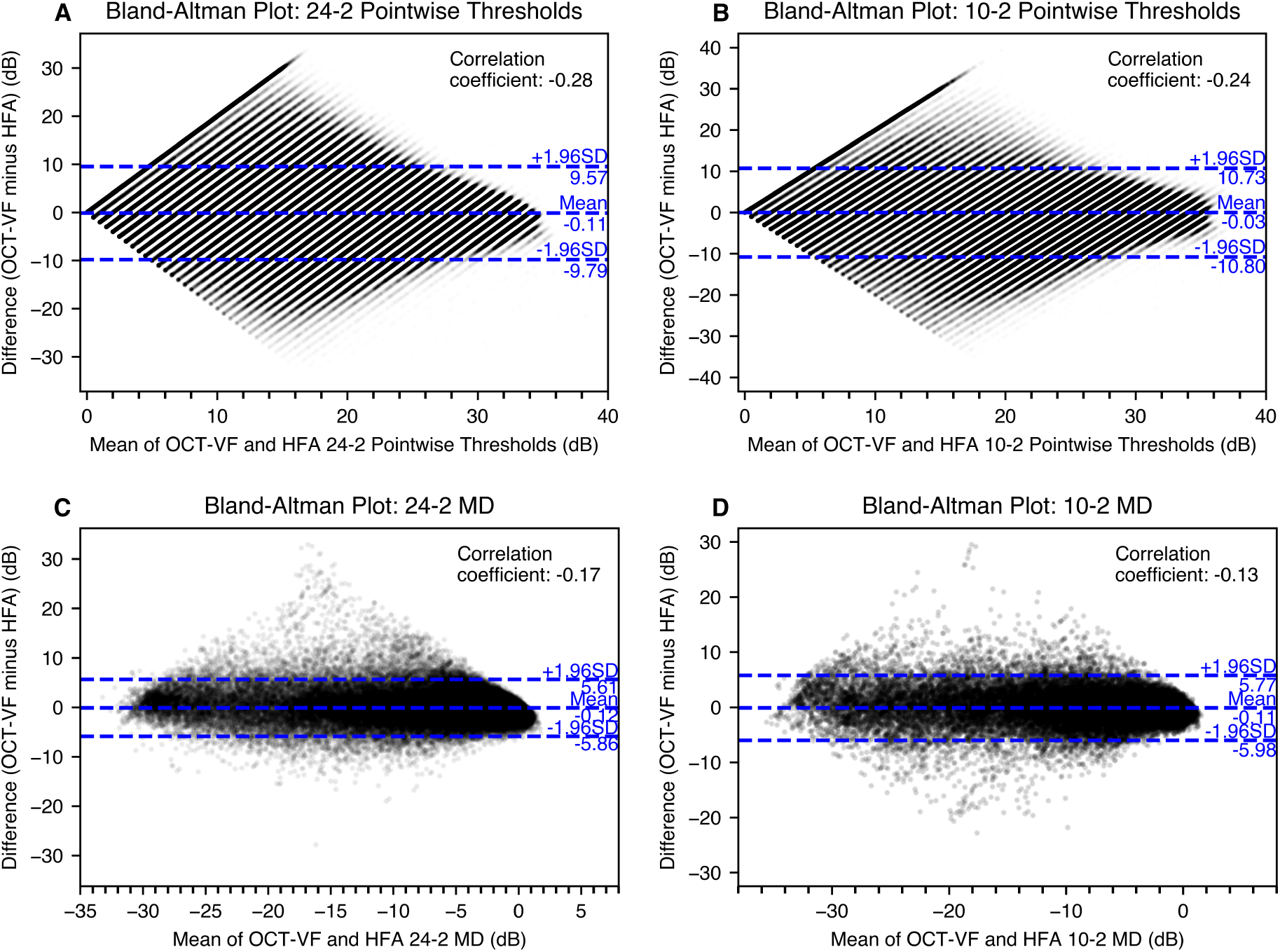
Bland-Altman plots for agreement between OCT-VF and HFA parameters. (A) 24-2 VF threshold (pointwise values), (B) 10-2 VF threshold (pointwise values), (C) 24-2 MD, and (D) 10-2 MD. The horizontal axis represents the mean of the OCT-VF and HFA values, whereas the vertical axis represents the difference between the OCT-VF and HFA values. The central dotted line indicates the mean difference, and the upper and lower dotted lines represent the 95% limits of agreement (±1.96 SD). The striped pattern observed for thresholds (A, B) arises from the discrete integer values of HFA measurements. The plots show the agreement between the OCT-VF and HFA values, with mean differences approaching zero for all analyses. Slight proportional biases were observed, with correlation coefficients ranging from -0.28 to -0.13. The magnitude of the error varied across different levels of VF loss. VF = visual field; MD = mean deviation; OCT = optical coherence tomography; OCT-VF = OCT-based estimated visual field; HFA = Humphrey Field Analyzer; SD = standard deviation.

**Table 1.** Training Dataset Characteristics.

| Patient characteristics | Value |
| --- | --- |
| Total number of patients | 13366 |
| Number of patients in HFA 24-2 | 12995 |
| Number of patients in HFA 10-2 | 4659 |
| Total number of eyes | 24313 |
| Number of eyes in HFA 24-2 | 23462 |
| Number of eyes in HFA 10-2 | 8085 |
| Total number of sets of paired data | 129007 |
| Number of sets of paired data in HFA 24-2 | 89711 |
| Number of sets of paired data in HFA 10-2 | 39296 |
| Number of HFA 24-2 tests per eye (mean ± SD) | 9.69 ± 7.61 |
| Number of HFA 10-2 tests per eye (mean ± SD) | 8.09 ± 5.14 |
| Proportion of SITA-Standard strategy in HFA 24-2 (%) | 46.5 |
| Proportion of SITA-Standard strategy in HFA 10-2 (%) | 47.2 |
| Number of OCT tests per eye (mean ± SD) | 5.31 ± 7.27 |
| Mean deviation of the HFA 24-2 (dB, mean ± SD) | -5.93 ± 7.65 |
| Mean deviation of the HFA 10-2 (dB, mean ± SD) | -8.03 ± 8.69 |
| Age (years, mean ± SD) | 65.0 ± 14.2 |
| SSI (mean ± SD) | 8.38 ± 1.03 |
| Focus (D, mean ± SD) | -2.60 ± 3.56 |

HFA, Humphrey Field Analyzer; SD, standard deviation; SITA, Swedish Interactive Threshold Algorithm; OCT, optical coherence tomography; SSI, signal strength index of OCT image.

**Table 2.** OCT-VF and HFA Parameter Correlations.

| Parameter | Pearson's r | Spearman's $\rho$ | MAE (dB) | ICC (2,1) |
| --- | --- | --- | --- | --- |
| 24-2 Thresholds | 0.863* | 0.819* | 3.15 ± 2.12 | 0.853 |
| 10-2 Thresholds | 0.881* | 0.832* | 3.45 ± 2.24 | 0.875 |
| 24-2 MD | 0.924* | 0.832* | 2.01 ± 2.13 | 0.922 |
| 10-2 MD | 0.939* | 0.909* | 2.10 ± 2.14 | 0.938 |

HFA = Humphrey Field Analyzer; OCT = optical coherence tomography; OCT-VF = OCT-based estimated visual field; MAE = mean absolute error; ICC = intraclass correlation coefficient; MD = mean deviation.
\* All correlations significant at $p < 0.001$

The model’s performance showed slight increases in error for severe cases (Fig. 5). The model’s estimation accuracy remained relatively stable across various OCT focus values, indicating the minimal impact of refractive errors on performance (Fig. 6). The spatial distribution of MAE across the 24-2 and 10-2 (Figs. 7–8) test patterns indicated that points in the nasal VF, where nerve fiber layer pathways are more extensively captured within the OCT scan area, tended to show lower error rates in severe cases. Conversely, points in the temporal VF closer to the optic disc tended to exhibit higher error rates in severe cases.

**Figure 5.**
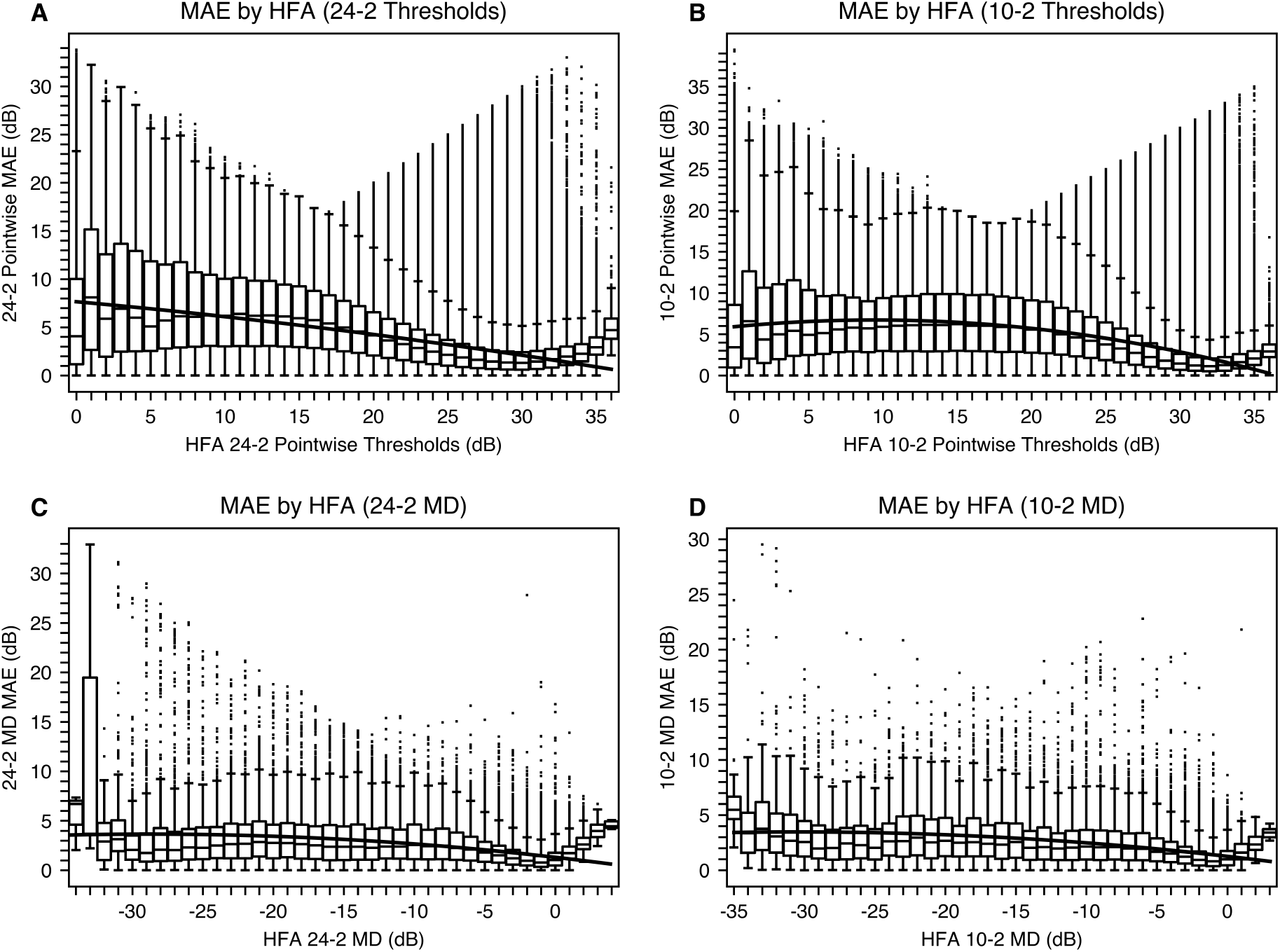
Relationships between VF severity and MAEs. (A) 24-2 VF threshold, (B) 10-2 VF threshold, (C) 24-2 MD, and (D) 10-2 MD. The horizontal axis represents VF severity, and the vertical axis represents MAE (dB). Each panel shows boxplots of MAEs along with a quadratic regression curve. For thresholds (A and B), there are slight increases in MAE in more severe cases. For MD (C and D), MAEs remain relatively stable across severity levels, with a notable exception at -33 dB in panel (C). Prior single-center analysis indicated that cases with extreme MD values and high estimation errors were often related to non-organic VF loss, technical measurement errors, or neurological visual pathway disorders rather than primary ocular pathologies. VF = visual field; MAE = mean absolute error; MD = mean deviation.

**Figure 6.**
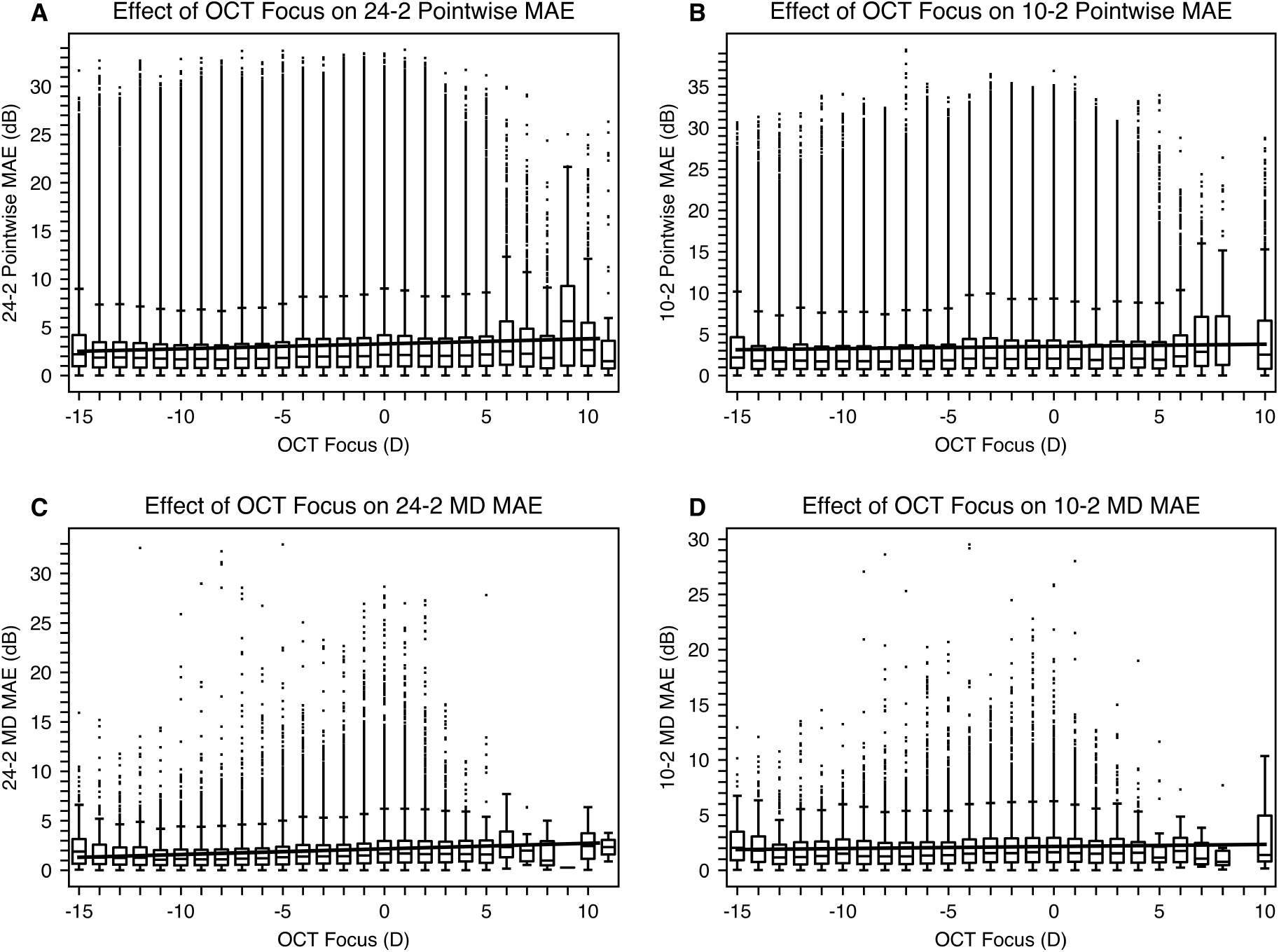
Impact of refractive error on model performance. (A) 24-2 VF thresholds, (B) 10-2 VF thresholds, (C) 24-2 MD, and (D) 10-2 MD. The horizontal axis represents OCT focus values (which closely correspond to refractive error), and the vertical axis represents MAE. Each panel shows boxplots of MAEs for each refractive error, with linear regression lines. Slight increases in MAEs are observed for hyperopic eyes across all measures. Linear regression analysis revealed slopes of 0.052 dB/D (24-2 VF thresholds), 0.027 dB/D (10-2 VF thresholds), 0.056 dB/D (24-2 MD), and 0.019 dB/D (10-2 MD). These minimal slopes quantify the negligible impact of refractive errors on model performance. OCT = optical coherence tomography; VF = visual field; MD = mean deviation; D = diopter; MAE = mean absolute error.

**Figure 7.**
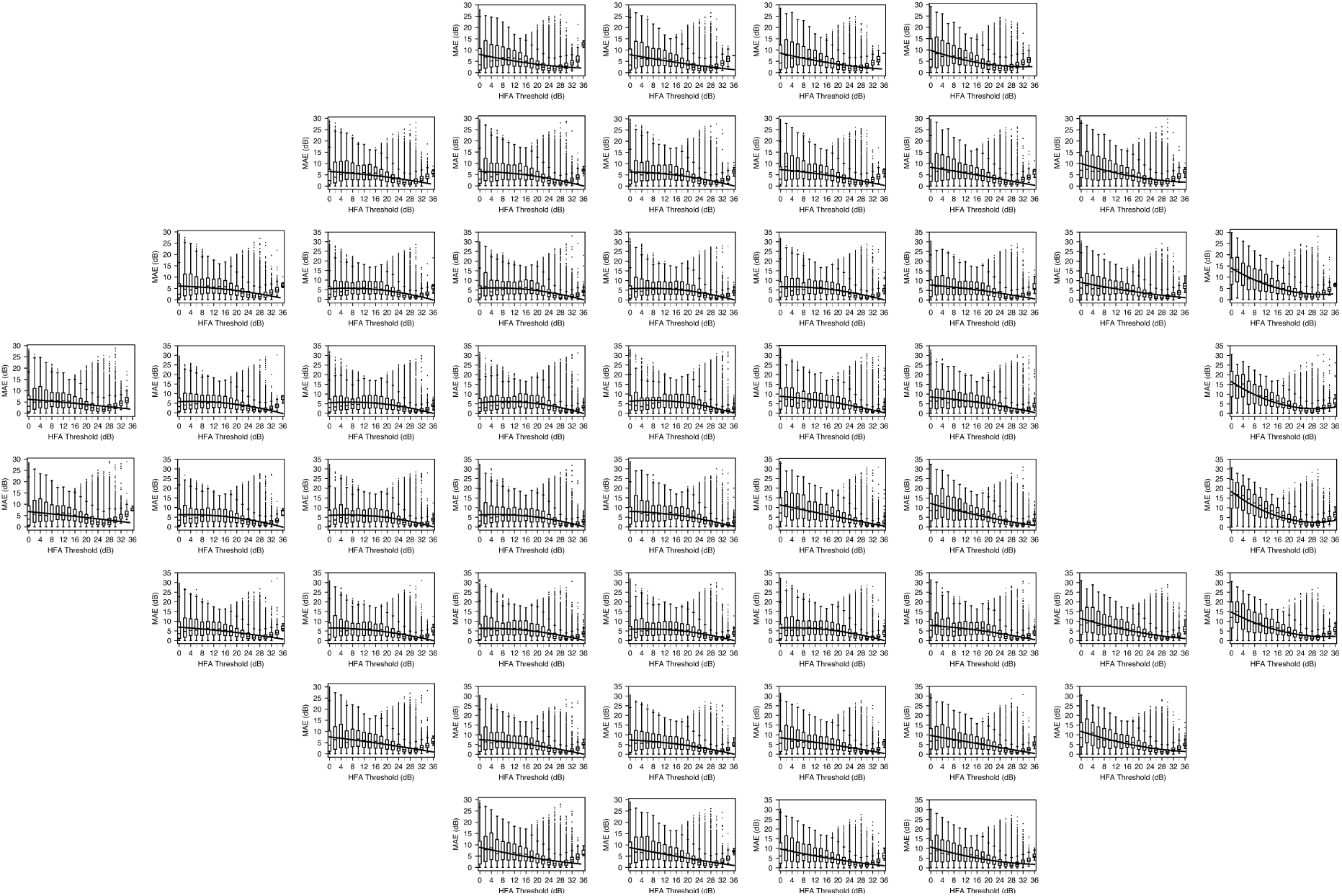
Spatial distribution of estimation accuracy across the 24-2 VF. The boxplots show MAE values for each test point, with quadratic regression curves showing the relationship between severity (horizontal axis) and MAE (vertical axis). Since the OCT imaging covered a 9 mm × 9 mm square centered on the macula (approximately extending to the optic disc), estimation accuracy varies across the VF. In severe cases, points in the nasal VF, where nerve fiber layer information is captured within the scan area, tended to show lower error rates. In contrast, points closer to the optic disc in the temporal VF tended to show higher error rates in severe cases, particularly in regions beyond the area covered by the OCT scan. This pattern suggests a relationship between OCT scan coverage and estimation accuracy across different VF regions. Data from left eyes were horizontally flipped and converted to right eye position for standardized analysis. MAE = mean absolute error; VF = visual field; OCT = optical coherence tomography.

**Figure 8.**
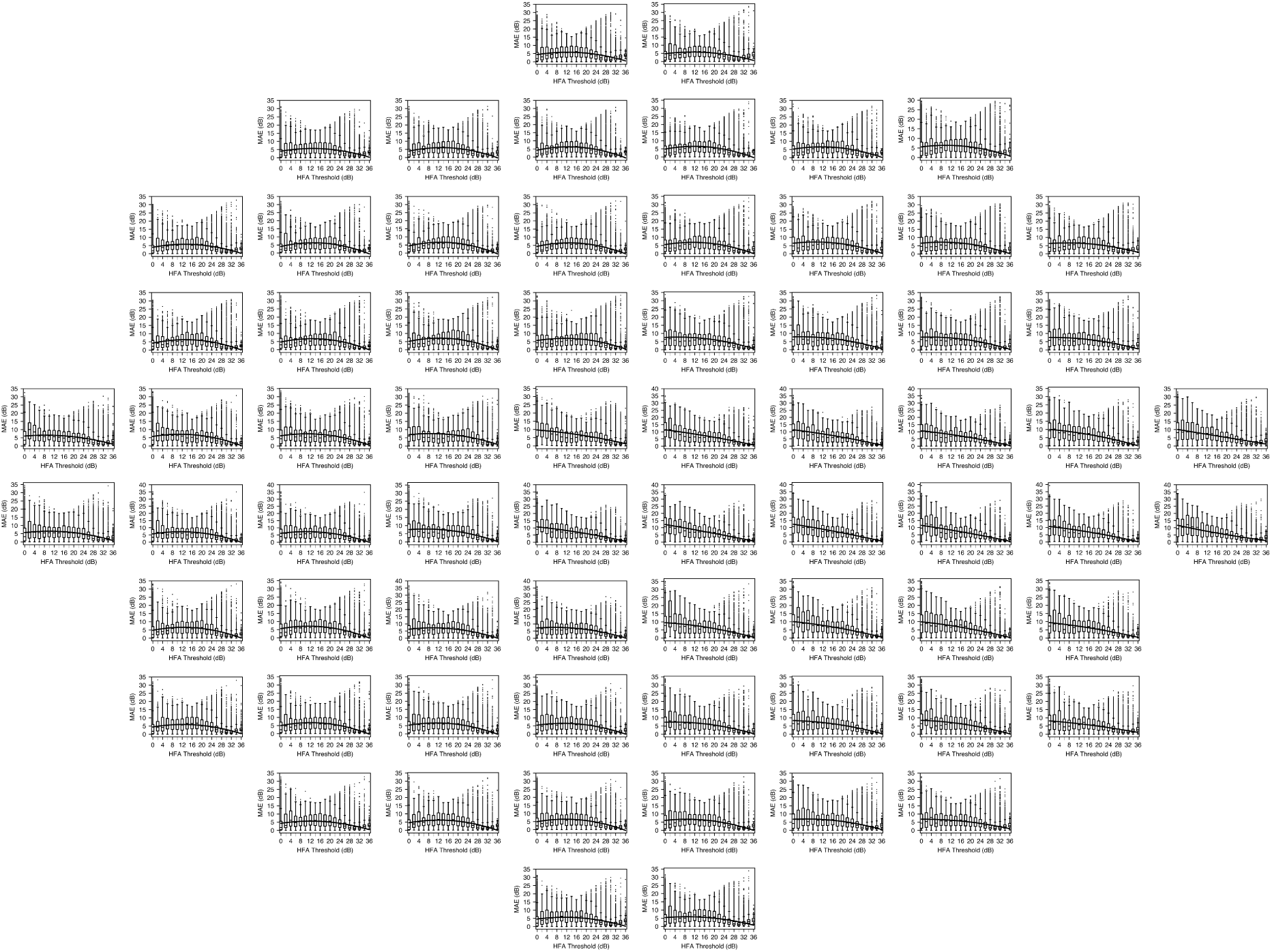
Spatial distribution of estimation accuracy across the 10-2 VF. Similar to Figure 7, these boxplots illustrate how MAE varies with threshold values across the central VF, with quadratic regression curves showing the relationship between severity (horizontal axis) and MAE (vertical axis). In severe cases, points in the nasal VF, where nerve fiber layer pathways are captured within the OCT scan area, tended to show lower error rates. Conversely, points in the temporal VF closer to the optic disc tended to show higher error rates in severe cases. This pattern suggests a relationship between neural pathway coverage in the OCT scan and estimation accuracy. Data from left eyes were horizontally flipped and converted to right eye position to ensure consistent spatial representation. MAE = mean absolute error; VF = visual field; OCT = optical coherence tomography.

## Longitudinal Dataset Characteristics for Comparative Analysis

For the longitudinal analysis comparing OCT-VF and HFA variability, we created independent time-series datasets for each method, as illustrated in Figure 9. Using jackknife resampling, we analyzed the deviations from regression lines for both OCT-VF and HFA measurements separately to determine which method demonstrated less variability and better progression detection capability. Table 3 summarizes the characteristics of these time-series datasets used for comparative analysis. After standardizing the number of measurements between groups, we observed that the HFA group had slightly longer mean observation periods (24-2: 2585 ± 1184 days, 10-2: 2355 ± 1089 days) than the OCT-VF group (24-2: 2572 ± 1191 days, 10-2: 2323 ± 1101 days; all p < 0.001, Wilcoxon signed-rank test). Because longer observation periods typically provide greater statistical power for detecting progression, this difference represents a conservative comparison condition that slightly favors the HFA group. A subanalysis including only SITA-Standard tests showed a higher proportion of more severe cases compared to the full dataset (Supplementary Table 4).

**Figure 9.**
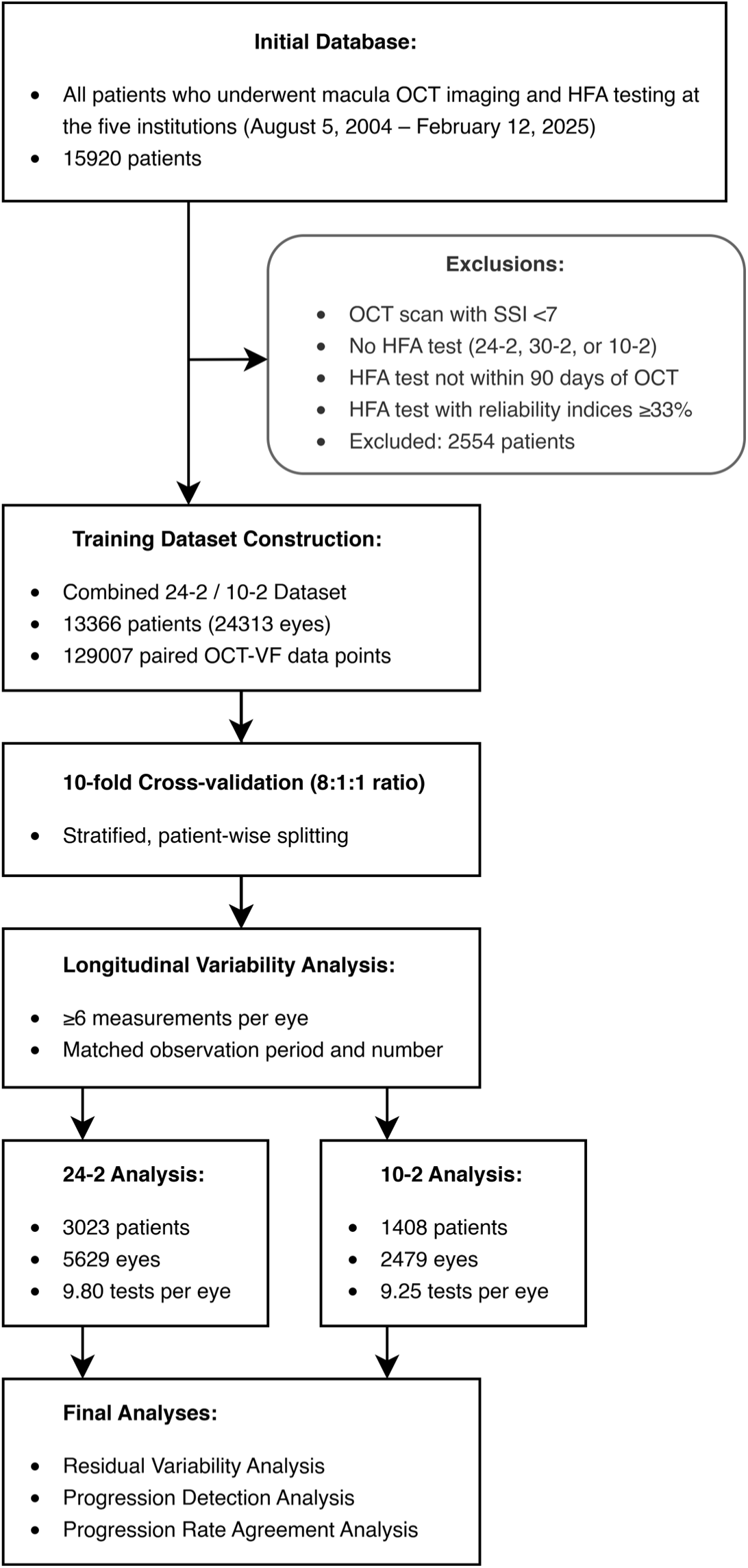
Study flowchart illustrating patient selection criteria and analytical workflow. Starting with 15,920 patients, exclusion criteria were applied, resulting in 13,366 patients included in the training dataset. After model development through 10-fold cross-validation, longitudinal variability analysis was performed on eyes with ≥6 measurements, with matched observation periods between OCT-VF and HFA groups. OCT = optical coherence tomography; HFA = Humphrey Field Analyzer; SSI = signal strength index; OCT-VF = OCT-based estimated visual field.

**Table 3.** OCT-VF and HFA Longitudinal Dataset Characteristics.

| Characteristics | OCT-VF<br>24-2 | HFA 24-2 | OCT-VF<br>10-2 | HFA 10-2 |
| --- | --- | --- | --- | --- |
| Number of patients | 3023 | 3023 | 1408 | 1408 |
| Number of eyes | 5629 | 5629 | 2479 | 2479 |
| Age (years) | 65.7 ± 13.4 | 65.2 ± 13.5 | 66.3 ± 13.3 | 65.9 ± 13.3 |
| Mean deviation (dB) | -6.39 ± 7.21* | -5.89 ± 7.42 | -8.43 ± 8.34* | -8.17 ± 8.53 |
| Number of tests | 9.80 ± 3.56 | 9.80 ± 3.56 | 9.25 ± 2.99 | 9.25 ± 2.99 |
| SITA-Standard (%) | - | 44.5 | - | 47.4 |
| Follow-up duration<br>(days) | 2572 ± 1191 | 2585 ± 1184 | 2323 ± 1101 | 2355 ± 1089 |

OCT = optical coherence tomography; OCT-VF = OCT-based estimated visual field; HFA = Humphrey Field Analyzer; SITA, Swedish Interactive Threshold Algorithm. The values are presented as the means ± standard deviations.
\*Mean deviations for OCT-VF are estimates derived from the OCT-VF models.

### Analysis of Visual Field Measurements at the Lower Threshold Limit

Figure 10 revealed that OCT-VF showed fewer measurements clustered at 0 dB compared to HFA, suggesting different behaviors in severely damaged VF regions. Longitudinal analysis (Fig. 11) confirmed that HFA measurements were more likely to persist at 0 dB, whereas OCT-VF values more often fluctuated slightly above the threshold floor. When recovery from 0 dB occurred, HFA values showed large and variable increases (24-2: 12.2 ± 8.2 dB; 10-2: 11.9 ± 8.7 dB), while OCT-VF changes remained small and stable (24-2: 1.7 ± 1.3 dB; 10-2: 1.9 ± 1.5 dB). SITA-Standard–only subanalyses confirmed these trends (Supplementary Figs. 12–13).

**Figure 10.**
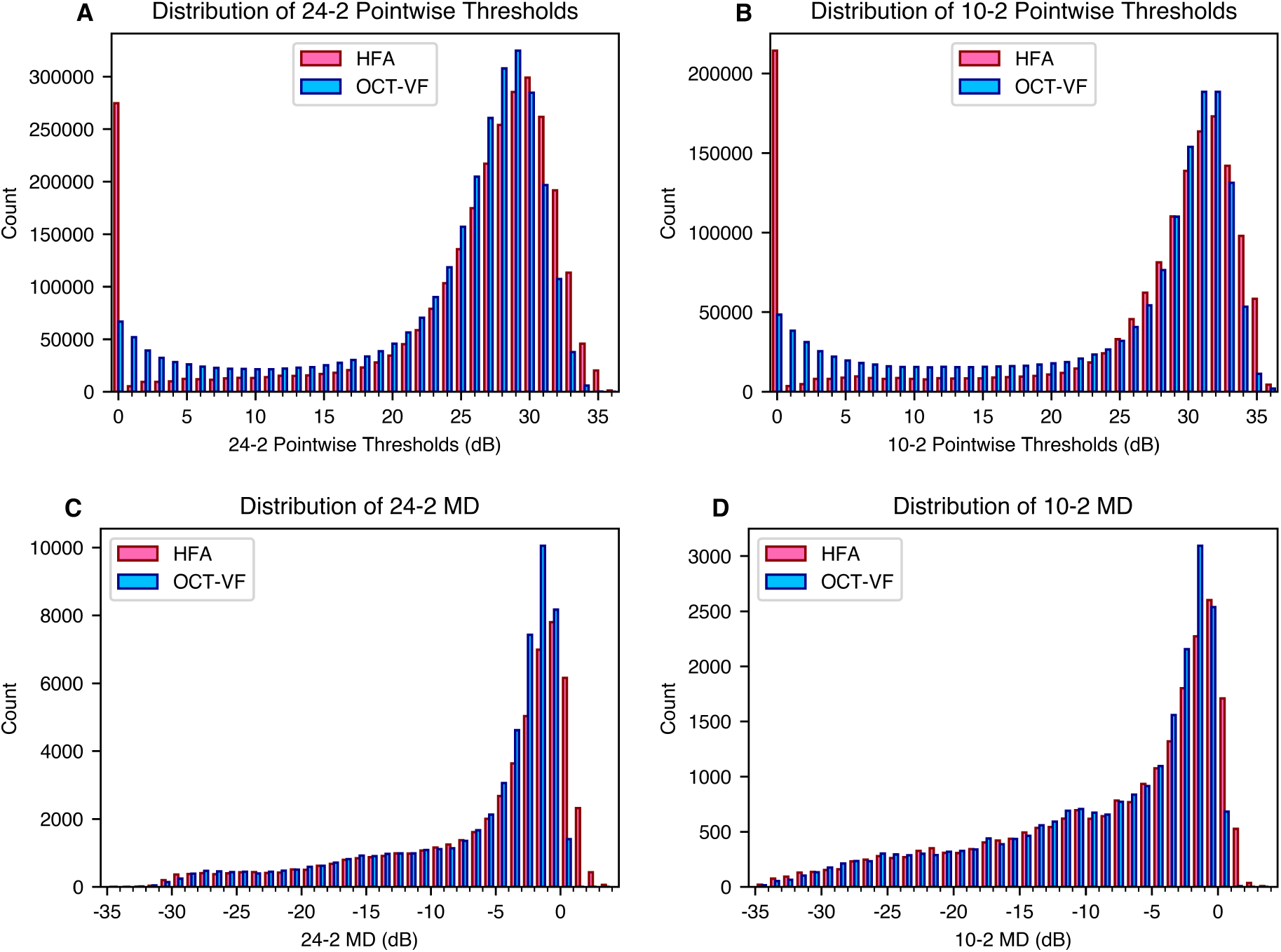
Population distribution by severity level. (A, B) show threshold distributions for 24-2 and 10-2 test patterns, respectively, while (C, D) show MD distributions for 24-2 and 10-2 patterns. In the threshold distributions (A, B), there is a notable discrepancy at 0 dB, with OCT-VF showing fewer measurements at this floor value compared to HFA. This difference prompted the further longitudinal analysis shown in Figure 11 to investigate the behavior of measurements following initial 0 dB readings. HFA = Humphrey Field Analyzer; OCT = optical coherence tomography; OCT-VF = OCT-based estimated visual field; MD = mean deviation.

**Figure 11.**
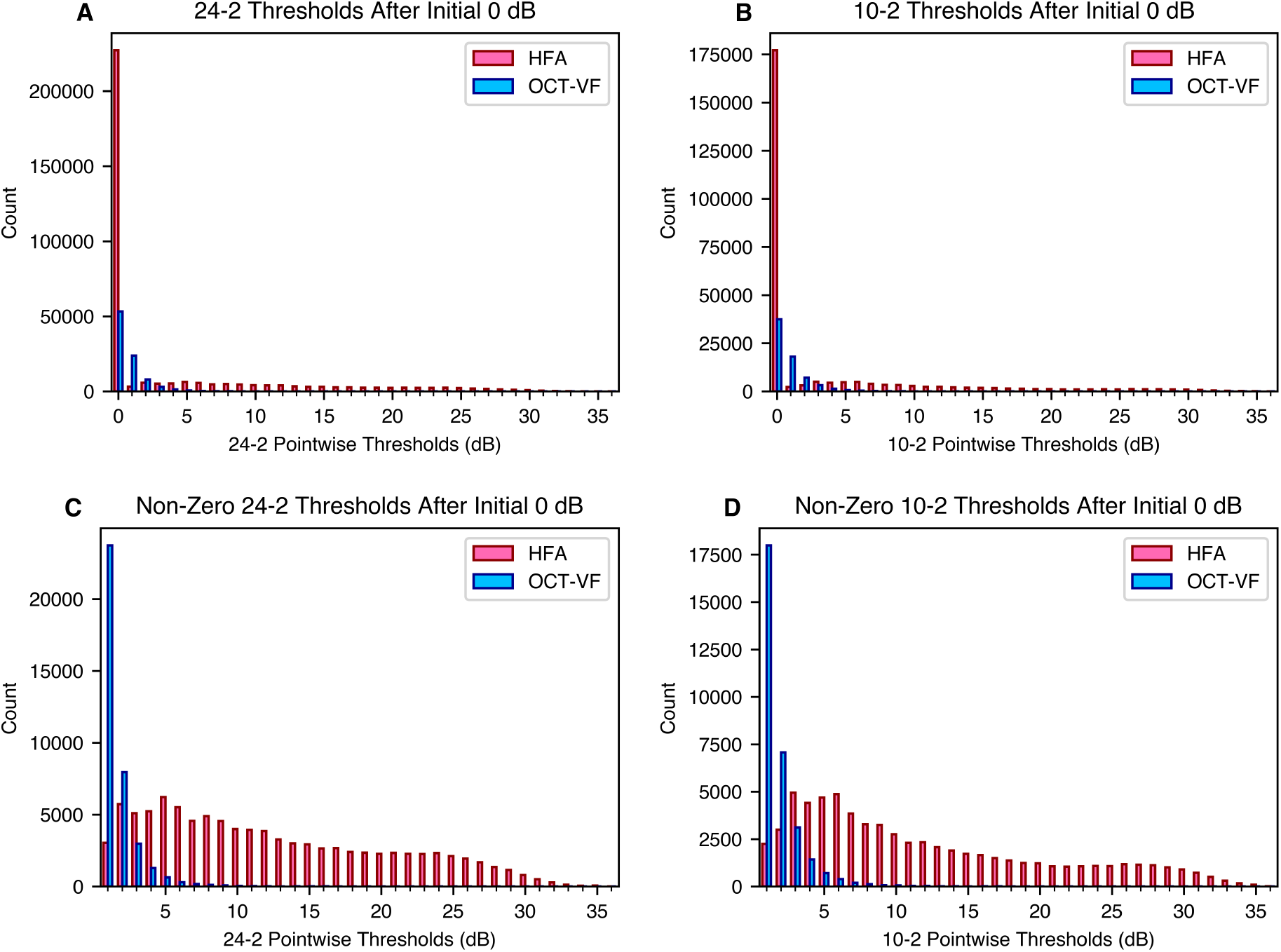
Longitudinal Analysis of Visual Field Thresholds Following Initial 0 dB Measurements. (A, B) show distributions of all pointwise thresholds following initial 0 dB readings in 24-2 and 10-2 tests. HFA measurements remain at 0 dB in 70.0% (24-2) and 72.5% (10-2) of cases, compared to 58.7% (24-2) and 54.5% (10-2) for OCT-VF. (C, D) show distributions of non-zero thresholds after initial 0 dB readings in 24-2 and 10-2 tests. HFA measurements showed substantial recovery (24-2: 12.2 ± 8.2 dB, 10-2: 11.9 ± 8.7 dB), while OCT-VF concentrate at lower values (24-2: 1.7 ± 1.3 dB, 10-2: 1.9 ± 1.5 dB). HFA = Humphrey Field Analyzer; OCT = optical coherence tomography; OCT-VF = OCT-based estimated visual field.

### Residual Variability Analysis using Generalized Estimating Equations (GEE)

Table 5 presents the residual variability and standard deviations for OCT-VF and HFA datasets. For both 24-2 and 10-2 test patterns, the OCT-VF group showed lower residual variability than the HFA group. For thresholds, OCT-VF vs. HFA residual variability were 0.89 vs. 2.24 dB for 24-2, and 1.06 vs. 2.36 dB for 10-2. For MDs, OCT-VF vs. HFA residual variability were 0.58 vs. 1.12 dB for 24-2, and 0.70 vs. 1.12 dB for 10-2. We compared the residual variability between the OCT-VF and HFA datasets using the GEE models described in the Methods section. OCT-VF exhibited significantly lower residual variability than HFA across all four analyses (p < 0.001), with all results remaining significant after Bonferroni correction (p < 0.0125). The SITA-Standard–only subanalysis confirmed these findings, with OCT-VF showing lower residual variability than HFA across all parameters (Supplementary Table 6).

**Table 5.** Residual Variability Comparison: OCT-VF vs. HFA.

| **Variability**<br>**Metric** | **OCT-VF**<br>**Mean ± SD**<br>**(dB)<sup>\*</sup>** | **HFA**<br>**Mean ± SD**<br>**(dB)<sup>\*</sup>** | **Difference<sup>†</sup>**<br>**(95% CI, dB)** | **P-value** | **Working**<br>**Correlation<sup>‡</sup>** |
| --- | --- | --- | --- | --- | --- |
| 24-2<br>Threshold | 0.89 ± 0.76 | 2.24 ± 2.06 | -1.36<br>(-1.39, -1.34) | < 0.001 | 0.42 |
| 10-2<br>Threshold | 1.06 ± 0.94 | 2.36 ± 2.46 | -1.30<br>(-1.34, -1.26) | < 0.001 | 0.38 |
| 24-2 MD | 0.58 ± 0.54 | 1.12 ± 0.92 | -0.54<br>(-0.57, -0.52) | < 0.001 | 0.32 |
| 10-2 MD | 0.70 ± 0.61 | 1.12 ± 1.00 | -0.42<br>(-0.46, -0.39) | < 0.001 | 0.37 |

MD = mean deviation; OCT = optical coherence tomography; OCT-VF = OCT-based estimated visual field; HFA = Humphrey Field Analyzer; CI = confidence interval.
<sup>\*</sup>The values for OCT-VF and HFA represent the residual variability calculated using jackknife resampling, measuring absolute deviations from the regression line for each method independently; lower values indicate less variability and better measurement consistency.
<sup>†</sup>The Difference column represents the difference between OCT-VF and HFA residual variability (OCT-VF minus HFA) calculated using generalized estimating equations (GEE), adjusting for follow-up duration, age, clustering by eye and patient, and measurement point location for threshold analyses only. Negative values indicate lower variability in OCT-VF.
<sup>‡</sup>Working Correlation represents the correlation between measurements from the same eye in the GEE model.

**Table 7.** Longitudinal Correlation Metrics: OCT-VF vs. HFA.

| **Metric** | **Test Type** | **Spearman's $\rho$**<br>**(Mean ± SD)** | **Significant**<br>**Events (n)<sup>\*</sup>** |
| --- | --- | --- | --- |
| 24-2 Clusters | OCT-VF | -0.372 ± 0.437 | 7582 / 56290 |
| 24-2 Clusters | HFA | -0.169 ± 0.455 | 2783 / 56290 |
| 10-2 Clusters | OCT-VF | -0.373 ± 0.442 | 2552 / 19832 |
| 10-2 Clusters | HFA | -0.249 ± 0.459 | 1409 / 19832 |
| 24-2 MD | OCT-VF | -0.459 ± 0.425 | 1638 / 5629 |
| 24-2 MD | HFA | -0.145 ± 0.515 | 770 / 5629 |
| 10-2 MD | OCT-VF | -0.489 ± 0.419 | 731 / 2479 |
| 10-2 MD | HFA | -0.284 ± 0.510 | 492 / 2479 |

HFA = Humphrey Field Analyzer; OCT = optical coherence tomography; MD = mean deviation; OCT-VF = OCT-based estimated visual field; SD = standard deviation.
<sup>\*</sup>Significant Events (n) are the "number of significant events / total number of observations." The numerator represents cases with statistically significant progression after Bonferroni correction ( $p < 0.0125$ ) for the four main parameters and additional Holm correction for clusters, using Spearman's rank correlation coefficient. The denominator represents the total number of cluster regions or eyes analyzed.

### Factors Influencing Residual Variability in OCT-VF and HFA Measurements

When examining the relationship between VF severity and residual variability (Fig. 14), quadratic regression analysis demonstrated that OCT-VF exhibited lower variability than HFA measurements across most severity levels for both 24-2 and 10-2 test patterns. This advantage was observed predominantly in threshold measurements and MD values for mild to moderate severity, with a slight reversal noted in severe cases for MD values.

**Figure 14.**
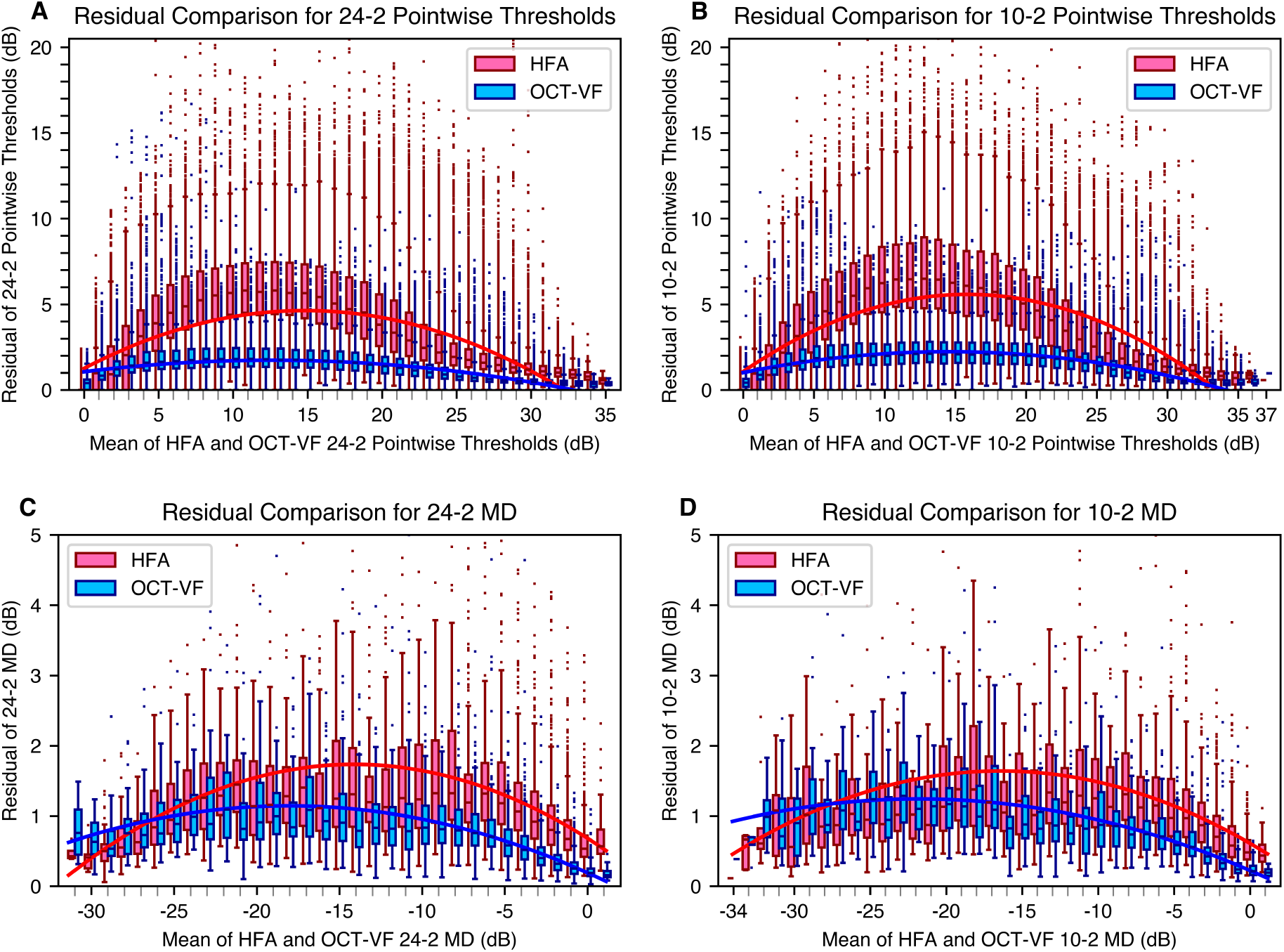
Comparison of residual variability between OCT-VF and HFA measurements across VF severity levels. (A) 24-2 thresholds (pointwise values), (B) 10-2 thresholds (pointwise values), (C) 24-2 MD, and (D) 10-2 MD. The horizontal axis represents VF severity, and the vertical axis represents residual variability. Each panel shows boxplots of residuals for each severity and quadratic regression lines. The quadratic regression lines for the OCT-VF dataset generally lie below those of the HFA dataset, indicating lower residual variability across most levels of VF severity for OCT-VF, though a slight reversal is observed in severe cases for both 24-2 and 10-2 MD. For each time point, the x-axis values represent the average of each measurement and its closest time-matched counterpart from the other modality. HFA = Humphrey Field Analyzer; OCT = optical coherence tomography; VF = visual field; OCT-VF = OCT-based estimated visual field; MD = mean deviation.

The spatial distribution of residual variability, visualized as heat maps, further confirmed this finding (Fig. 15). OCT-VF demonstrated substantially lower variability across all test points compared to HFA, with values ranging from 0.72 to 1.25 dB for OCT-VF versus 1.76 to 3.04 dB for HFA, representing a mean variability reduction of 60.4% for 24-2 and 55.1% for 10-2 test patterns.

**Figure 15.**
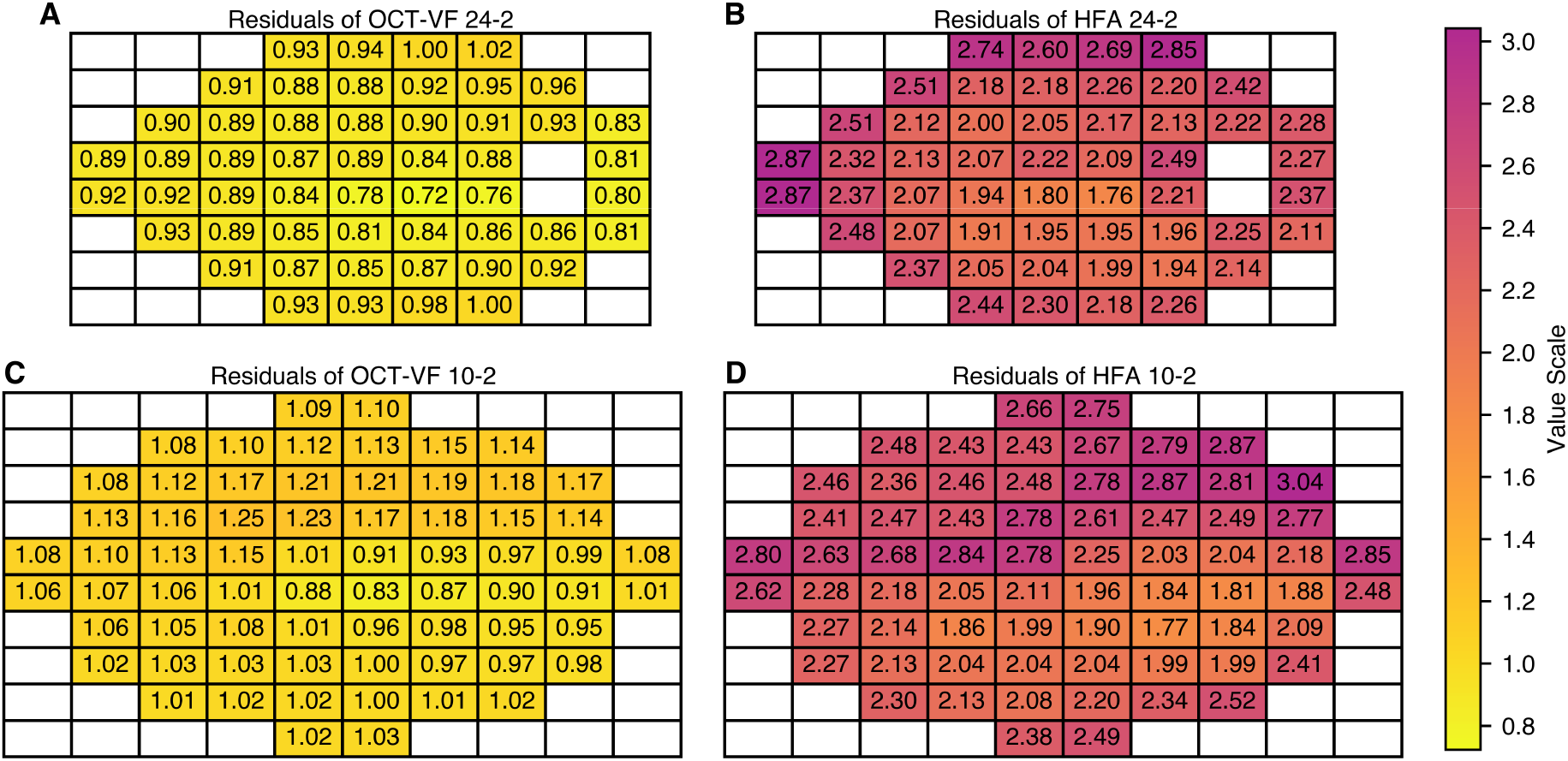
Heat maps of residual variability for each test point. (A) OCT-VF 24-2, (B) HFA 24-2, (C) OCT-VF 10-2, and (D) HFA 10-2. The color scale represents the magnitude of residual variability at each test point. OCT-VF shows consistently lower residual variability than HFA measurements across all test points, with values ranging from 0.72 to 1.25 dB for OCT-VF compared to 1.76 to 3.04 dB for HFA measurements. We horizontally flipped the left eye data and integrated them with the right eye data. OCT = optical coherence tomography; OCT-VF = OCT-based estimated visual field; HFA = Humphrey Field Analyzer.

Age-related analysis (Fig. 16) revealed that while both methods showed increased variability with advancing age, OCT-VF maintained its advantage across all age groups. The difference in variability between methods widened in older patients, with OCT-VF showing a less pronounced age-related increase in variability.

**Figure 16.**
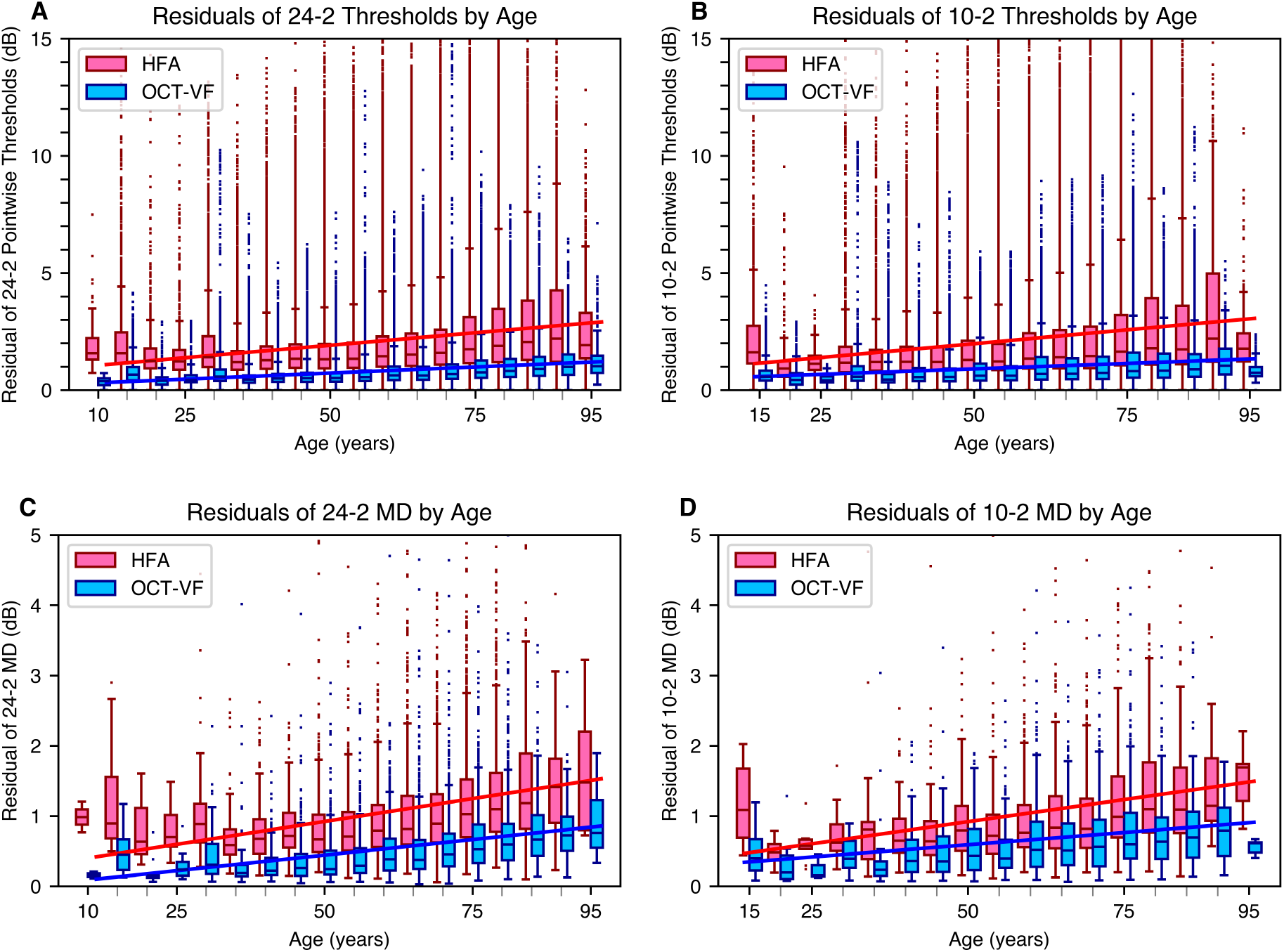
Comparison of residual variability between HFA measurements and OCT-VF across age groups. (A) 24-2 thresholds, (B) 10-2 thresholds, (C) 24-2 MD, and (D) 10-2 MD. The horizontal axis represents age in 5-year increments, and the vertical axis represents residual variability. Each panel shows boxplots of residuals for each age group, along with linear regression lines for HFA (red) and OCT-VF (blue). The regression lines for OCT-VF consistently lie below those for HFA, indicating lower residual variability across all age groups. Both methods show increased variability with age, but HFA demonstrates steeper slopes, particularly for threshold measurements. The slopes of the regression lines (in dB/year) for HFA and OCT-VF are (A) 0.0213 vs. 0.0105, (B) 0.0236 vs. 0.0094, (C) 0.0130 vs. 0.0089, and (D) 0.0126 vs. 0.0070, respectively. These values suggest that, compared with HFA, OCT-VF is less affected by age-related increases in variability. HFA = Humphrey Field Analyzer; OCT = optical coherence tomography; OCT-VF = OCT-based estimated visual field; MD = mean deviation.

### Progression Rate Analysis

OCT-VF demonstrated stronger correlations and detected more significant progression events than HFA across all parameters: 172% more for 24-2 clusters, 81.1% more for 10-2 clusters, 113% more for 24-2 MD, and 48.6% more for 10-2 MD. Bland-Altman analysis (Fig. 17) showed agreement in progression rates between methods, with mean differences (all ≤0.22 dB/year) and correlation coefficients (all ≤0.31). Boxplot analysis (Fig. 18) showed correlations between OCT-VF and HFA progression rates (Pearson’s r: 24-2 clusters 0.838, 10-2 clusters 0.846, 24-2 MD 0.831, 10-2 MD 0.863; all p < 0.001).

**Figure 17.**
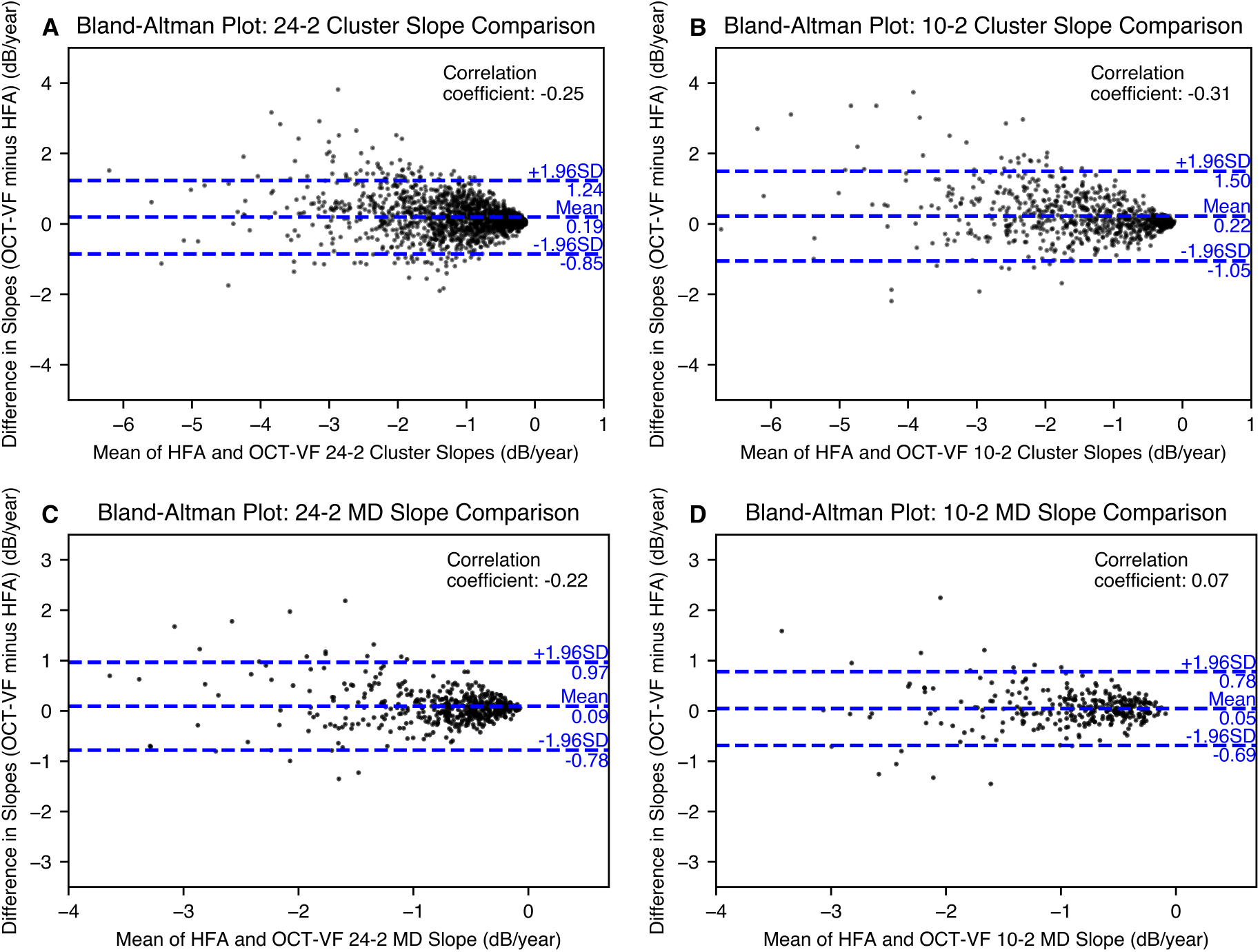
Bland-Altman plots comparing progression rates between OCT-VF and HFA. (A) 24-2 clusters, (B) 10-2 clusters, (C) 24-2 MD, and (D) 10-2 MD. The analysis includes only clusters or MDs that showed significant progression (after Bonferroni and Holm corrections) in both methods. The total number of progression events was 1635 in (A), 753 in (B), 517 in (C), and 315 in (D). Mean differences across all analyses were 0.22 dB/year or less, with correlation coefficients 0.31 or less. HFA = Humphrey Field Analyzer; OCT = optical coherence tomography; OCT-VF = OCT-based estimated visual field; MD = mean deviation.

**Figure 18.**
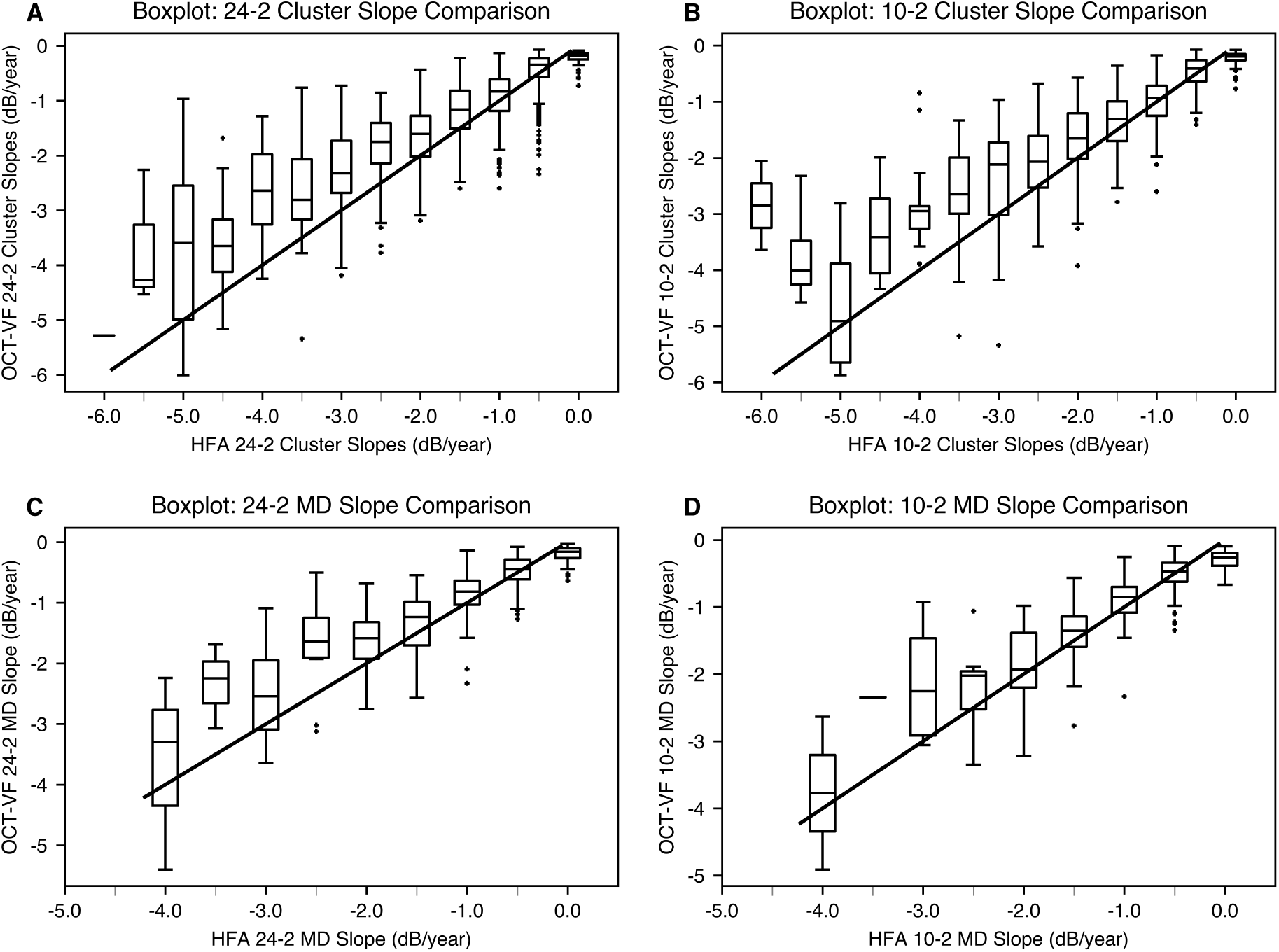
Boxplots comparing progression rates between HFA and OCT-VF. (A) 24-2 clusters, (B) 10-2 clusters, (C) 24-2 MD, and (D) 10-2 MD. The analysis includes only clusters or MDs that showed significant progression in both methods (2134, 849, 682, and 366 events, respectively). Each boxplot represents OCT-VF progression rates for the corresponding HFA progression rate. The solid line represents the identity line (x=y). Pearson’s correlation coefficients were 0.838 (A), 0.846 (B), 0.831 (C), and 0.863 (D), all p < 0.001. HFA = Humphrey Field Analyzer; OCT = optical coherence tomography; OCT-VF = OCT-based estimated visual field; MD = mean deviation.

## Discussion

This study demonstrated that our segmentation-free 3DCNN model objectively estimated VF with significantly lower residual variability than HFA measurements. The neural network likely converges toward the underlying "true" VF by averaging out random variability in individual HFA measurements—a key advantage of data-driven approaches in extracting stable patterns from noisy training data. Such convergence, however, is critically dependent on access to large-scale, diverse datasets. By leveraging a multicenter dataset of over 129,000 OCT-VF pairs without disease-specific exclusions, our model was able to generalize across a wide spectrum of ocular conditions and patient characteristics, thereby enhancing its robustness and clinical applicability.

Our analysis at the lower threshold limit (0 dB) revealed key differences between the methods. One contributing factor to the differing 0 dB counts between groups is that OCT-VF inherently produces predictions biased toward the central tendency in the presence of uncertainty. Another key difference was a potential threshold clipping effect in HFA: when true VF values fluctuate near 0 dB, HFA measurements are constrained to move upward, artificially accumulating 0 dB readings. This becomes apparent when examining how the two methods behaved when recovering from 0 dB readings. HFA values showed large and variable increases (∼12 dB), consistent with Gardiner et al.’s findings on reduced reliability below 19 dB.^19^ In contrast, OCT-VF changes remained small (∼2 dB), indicating greater stability. Although previous studies typically used HFA measurements as the reference standard because determining the true underlying VF is inherently difficult,^7–15^ these findings suggest that the conventional assumption of HFA measurements as the true reference standard may need reconsideration, and that OCT-based estimations might provide a more stable reflection of underlying visual function in certain clinical scenarios, particularly in areas with reduced sensitivity.

We employed a balanced methodological approach in two stages. First, we used HFA as a reference to establish baseline performance metrics for our OCT-VF model. Then, we independently analyzed longitudinal data for each method for our primary analysis of variability and progression detection. This approach allowed us to evaluate the intrinsic variability of both methods without assuming either as a gold standard.

OCT-based VF estimations demonstrated significantly lower residual variability than HFA measurements across all parameters. GEE analysis revealed OCT-VF’s superior consistency for 24-2 thresholds, 10-2 thresholds, 24-2 MD, and 10-2 MD, even after adjusting for confounding factors. This enhanced reliability persisted across regions, age groups, and most severity levels, with pronounced advantages in older populations. The progression analysis showed that OCT-VF identified more significant progression events despite the HFA group having longer observational periods.

Our findings have significant clinical implications. OCT-VF’s lower variability addresses a critical clinical challenge: distinguishing true disease progression from inherent test variability. The substantially lower residual variability provides increased statistical power and offers practical benefits, including improved efficiency in disease monitoring, more timely therapeutic interventions, potentially reduced patient burden, and lower healthcare costs. Our approach is intended to complement—not replace—standard automated perimetry, creating a synergistic relationship that leverages the strengths of both methods to enhance clinical decision-making. This objective approach may benefit patients with conditions that affect traditional VF testing reliability, such as limited test proficiency, advanced age, or cognitive impairments.^20^ Recent work by Mohammadzadeh et al. demonstrated that structural OCT measurements can also be valuable for predicting future functional glaucoma progression using deep learning, further highlighting the rich diagnostic information in OCT scans beyond their traditional structural applications.^21^

While our OCT-VF approach offers advantages, this study has several limitations. The model’s architecture and training process may introduce inherent biases, particularly in cases where structural OCT findings and functional visual field results are discordant. Additionally, our study was limited by using OCT devices from a single manufacturer with a specific imaging protocol. Future research should validate these findings through external studies with diverse populations, assess model performance across different disease subtypes, and evaluate compatibility with various OCT platforms.

## Conclusions

Our findings demonstrate that a segmentation-free, OCT-based 3DCNN model provides more consistent visual field estimations than standard automated perimetry. Rather than replacing HFA, this approach is intended as a complementary tool—leveraging routinely acquired OCT scans to offer objective, structure-based assessments using existing clinical data. The reduced variability of OCT-VF enhances statistical power for detecting progression across various ocular conditions. This more stable and objective assessment may facilitate earlier detection of true visual function changes, potentially enabling more timely therapeutic interventions and better preservation of vision, particularly in patients with conditions affecting conventional test reliability.

## Data Availability

We are unable to make the datasets publicly available due to privacy and ethical considerations related to patient data. We conducted the study on an opt-out basis, without obtaining explicit consent from all participants to release their raw data.

## Data availability statement

The datasets generated and/or analyzed during the current study are not publicly available due to privacy and ethical considerations. The study was conducted on an opt-out basis, and explicit consent for data sharing was not obtained from all participants.

## Author’s Contributions

**Makoto Koyama:** Conceptualization, Methodology, Software, Validation, Formal analysis, Investigation, Resources, Data Curation, Writing – Original Draft, Visualization, Project administration. **Hidenori Takahashi:** Supervision, Formal analysis, Validation, Investigation, Writing – Review & Editing. **Satoru Inoda:** Validation, Writing – Review & Editing. **Chihiro Mayama:** Investigation, Writing – Review & Editing**. Yuta Ueno:** Validation, Writing – Review & Editing. **Yoshikazu Ito:** Writing – Review & Editing. **Tetsuro Oshika:** Supervision, Writing – Review & Editing. **Masaki Tanito:** Supervision, Investigation, Writing – Review & Editing.

