## Supplementary Figure 12 for "OCT-based Visual Field Estimation via Segmentation-free 3D CNNs Shows Lower Longitudinal Variability than Standard Automated Perimetry"

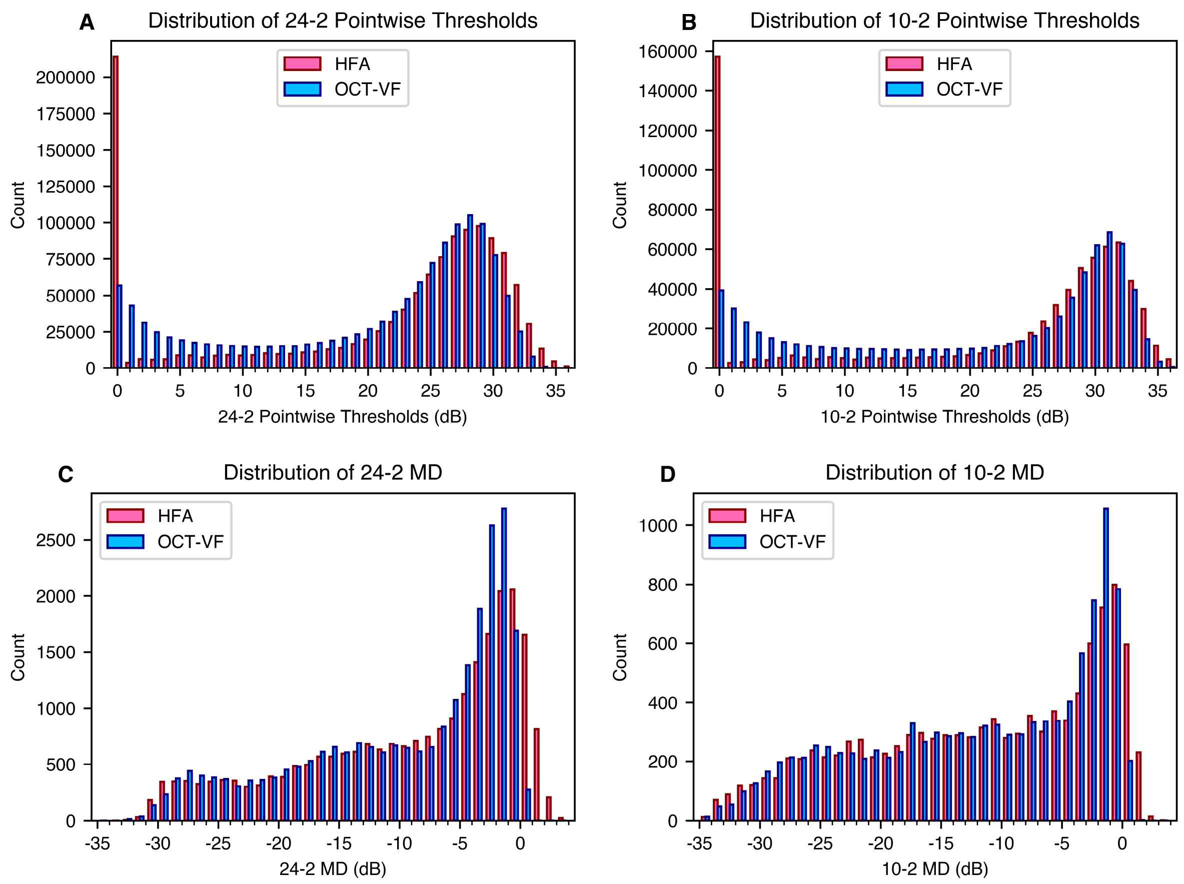


**Supplementary Figure 12.** SITA-Standard–Only: Population distribution by severity level. (A, B) show threshold distributions for 24-2 and 10-2 test patterns, respectively, while (C, D) show MD distributions for 24-2 and 10-2 patterns. In the threshold distributions (A, B), there is a notable discrepancy at 0 dB, with OCT-VF showing fewer measurements at this floor value compared to HFA. HFA = Humphrey Field Analyzer; OCT = optical coherence tomography; OCT-VF = OCT-based estimated visual field; MD = mean deviation; SITA, Swedish Interactive Threshold Algorithm.
