## Supplementary Figure 13 for "OCT-based Visual Field Estimation via Segmentation-free 3D CNNs Shows Lower Longitudinal Variability than Standard Automated Perimetry"

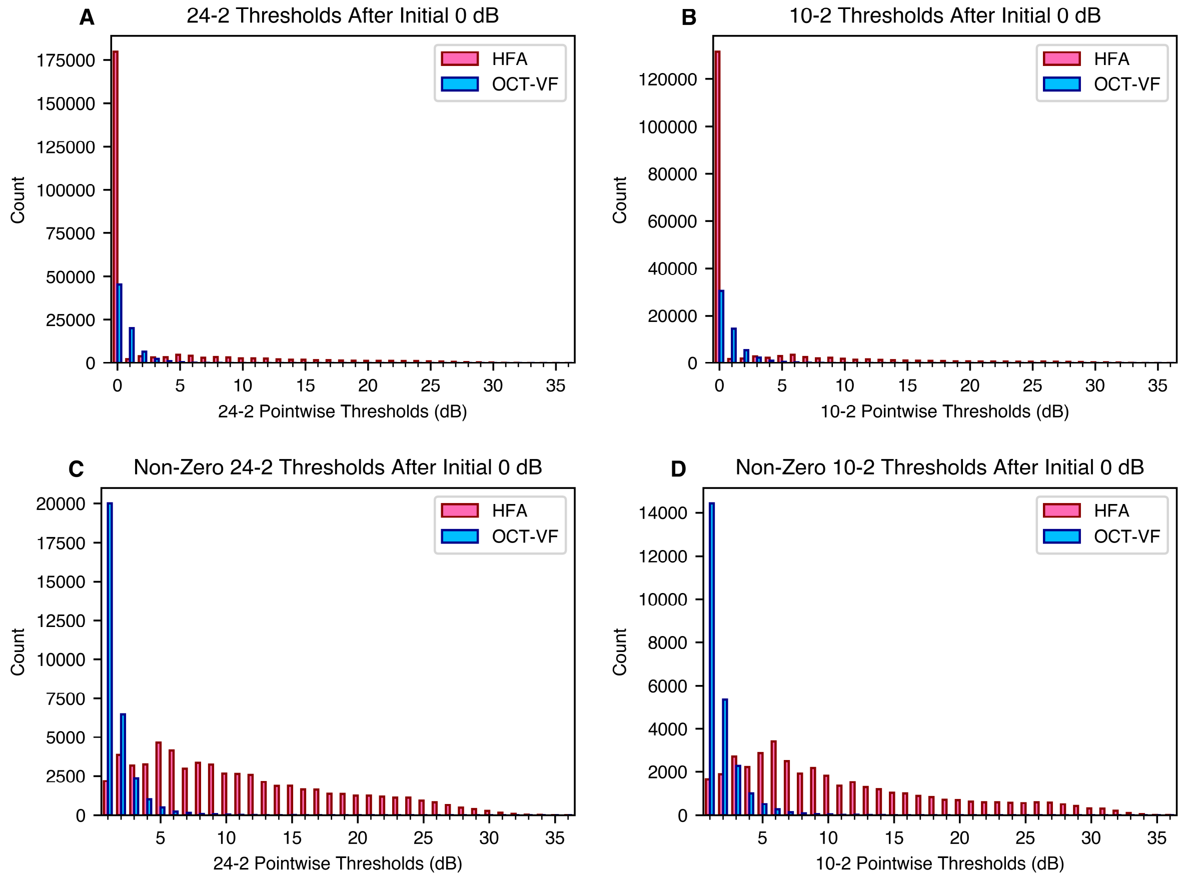


**Supplementary Figure 13.** SITA-Standard–Only: Longitudinal Analysis of Visual Field Thresholds Following Initial 0 dB Measurements.

(A, B) show distributions of all pointwise thresholds following initial 0 dB readings in 24-2 and 10-2 tests. HFA measurements remain at 0 dB in 74.9% (24-2) and 76.8% (10-2) of cases, compared to 59.4% (24-2) and 55.8% (10-2) for OCT-VF.

(C, D) show distributions of non-zero thresholds after initial 0 dB readings in 24-2 and 10-2 tests. HFA measurements showed substantial recovery (24-2: 11.1 ± 7.5 dB, 10-2: 11.2 ± 8.2 dB), while OCT-VF concentrate at lower values (24-2: 1.7 ± 1.3 dB, 10-2: 1.8 ± 1.5 dB).

SITA, Swedish Interactive Threshold Algorithm; HFA = Humphrey Field Analyzer; OCT = optical coherence tomography; OCT-VF = OCT-based estimated visual field.
