## Supplementary Table 4 for "OCT-based Visual Field Estimation via Segmentation-free 3D CNNs Shows Lower Longitudinal Variability than Standard Automated Perimetry"

**Supplementary Table 4.** SITA-Standard–Only: OCT-VF and HFA Longitudinal Dataset Characteristics

| Characteristics | OCT-VF 24-2 | HFA 24-2 | OCT-VF 10-2 | HFA 10-2 |
|---|---|---|---|---|
| Number of patients | 1385 | 1385 | 673 | 673 |
| Number of eyes | 2535 | 2535 | 1214 | 1214 |
| Age (years) | 67.8 ± 13.6 | 65.8 ± 13.6 | 67.3 ± 13.6 | 67.0 ± 13.5 |
| Mean deviation (dB) | -9.56 ± 8.39* | -8.99 ± 8.65 | -11.5 ± 9.42* | -11.4 ± 9.59 |
| Number of tests | 9.53 ± 3.61 | 9.53 ± 3.61 | 8.84 ± 2.61 | 8.84 ± 2.61 |
| Follow-up duration (days) | 2129 ± 1030 | 2210 ± 1087 | 1828 ± 803 | 1875 ± 775 |

OCT = optical coherence tomography; OCT-VF = OCT-based estimated visual field; HFA = Humphrey Field Analyzer; SITA, Swedish Interactive Threshold Algorithm. The values are presented as the means ± standard deviations.

*Mean deviations for OCT-VF are estimates derived from the OCT-VF models.
