## Supplementary Table 6 for "OCT-based Visual Field Estimation via Segmentation-free 3D CNNs Shows Lower Longitudinal Variability than Standard Automated Perimetry"

**Supplementary Table 6.** SITA-Standard–Only: Residual Variability Comparison

| Variability Metric | OCT-VF Mean ± SD (dB)* | HFA Mean ± SD (dB)* | Difference† (95% CI, dB) | Working Correlation‡ |
|---|---|---|---|---|
| 24-2 Threshold | 1.08 ± 0.87 | 2.48 ± 2.28 | -1.41 (-1.45, -1.37) | 0.42 |
| 10-2 Threshold | 1.18 ± 0.98 | 2.49 ± 2.57 | -1.30 (-1.37, -1.24) | 0.39 |
| 24-2 MD | 0.74 ± 0.63 | 1.25 ± 1.00 | -0.52 (-0.55, -0.48) | 0.34 |
| 10-2 MD | 0.80 ± 0.65 | 1.19 ± 1.12 | -0.38 (-0.44, -0.32) | 0.36 |

SITA, Swedish Interactive Threshold Algorithm; MD = mean deviation; OCT = optical coherence tomography; OCT-VF = OCT-based estimated visual field; HFA = Humphrey Field Analyzer; SD = Standard Deviation; CI = confidence interval.

*The values for HFA and OCT-VF represent the residual variability calculated using jackknife resampling, measuring deviations from the regression line for each method independently; lower values indicate less variability and better measurement consistency.

†The Difference column represents the OCT-VF minus the HFA value, with negative values indicating lower variability in OCT-VF.

‡Working Correlation represents the correlation between measurements from the same eye in the generalized estimating equations model.
